# Pediatric antibiotic use associated with respiratory syncytial virus and influenza in the United States, 2008-2018

**DOI:** 10.1101/2025.03.25.25324633

**Authors:** Laura M King, Katia J Bruxvoort, Sara Y Tartof, Joseph A Lewnard

## Abstract

**Background:** Understanding of the contributions of respiratory syncytial virus (RSV) and influenza infections to pediatric antibiotic use is limited. We aimed to estimate the proportions and incidence of outpatient antibiotic prescriptions associated with RSV and influenza infections in a sample of commercially-insured US children.

**Methods:** We conducted a retrospective study of outpatient antibiotic prescriptions dispensed to children in the Optum Clinformatics™ DataMart from 2008-2018. We used negative binomial time-series models regressing weekly antibiotic prescriptions against RSV and influenza circulation measures to estimate counterfactual rates of antibiotic prescriptions in the presence and absence of RSV and influenza circulation overall, by age group, census division, and antibiotic class. We considered both syndromic (medical claims) and laboratory (National Respiratory and Enteric Virus Surveillance System) RSV and influenza measures and controlled for age, division, 13-valent pneumococcal conjugate vaccine introduction, and seasonal and secular trends.

**Results:** An estimated 6.3% (95% confidence interval 5.2-7.3%) and 3.4% (3.1-3.8%) of antibiotic prescriptions were associated with RSV and influenza, respectively. These estimates translate to 72.6 (59.7-85.9) RSV-associated and 40.0 (35.1-45.1) influenza-associated antibiotic prescriptions per 1000 children annually. RSV-associated antibiotic prescription incidence was highest among children aged :::5 years while influenza-associated antibiotic prescriptions were highest among children >5 years. Macrolides were the antibiotic class for which RSV and influenza accounted for the greatest share of prescribing.

**Conclusions:** RSV and influenza account for meaningful proportions of pediatric antibiotic prescriptions. Measures to prevent RSV and influenza infections in children, including immunization, may reduce antibiotic use and aid in mitigating antibiotic resistance.

## BACKGROUND

Reducing outpatient antibiotic use is a cornerstone of strategies to prevent antimicrobial resistance.^1,2^ Acute respiratory infections (ARIs) are leading causes of pediatric outpatient antibiotic use: in 2014-15, ARIs accounted for 62% of outpatient antibiotic prescriptions for children from doctors’ offices and emergency departments.^3^ Respiratory syncytial virus (RSV) and influenza are common causes of ARI.^4–6^ Although frequently noted as etiologies in lower respiratory tract infections (LRTI), RSV and influenza also cause other ARIs (e.g., acute otitis media [AOM], pharyngitis), independently and via synergistic interactions with other pathogens.^7^ RSV and influenza infections may lead to antibiotic use due to diagnostic uncertainty in outpatient-managed ARIs, inappropriate prescribing, and treatment for concomitant bacterial infections.

Despite contributions of RSV and influenza in ARIs, evidence quantifying their role in outpatient antibiotic prescribing to US children remains limited. Vaccination against influenza has been found to reduce antibiotic use in randomized^8^ and observational studies.^9,10^ In addition, maternal RSV vaccination reduced antibiotic use among infants over the first three months of life in a randomized trial of a RSV vaccine candidate.^11^ However, identifying the proportion of all pediatric antibiotic use attributable to these pathogens poses challenges, in part due to sparse diagnostic testing in outpatient settings, non-specific respiratory manifestations, and diagnostic coding practices. Time-series analyses leveraging distinct seasonal pathogen circulation patterns provide a basis for quantifying antibiotic utilization associated with these infections.^12–14^ For instance, a time-series study of Scottish children found that RSV and influenza accounted for 6.9% and 2.4%, respectively, of all antibiotic prescriptions among children aged <5 years.^12^ Using similar methods, a study of children enrolled in Kaiser Permanente Northern California from 2010-2018 found that 1.4-2.7% of antibiotic prescriptions were associated with influenza.^13^ However, generalizability of these findings to the broader US pediatric population is limited given variation in prescribing patterns across healthcare systems, geographic regions^15,16^ and countries.^17^

Understanding RSV and influenza contributions to antibiotic use may guide prescribing practices and inform assessments of vaccination impacts on antibiotic use and antimicrobial resistance.^18^ We therefore aimed to estimate the proportions and incidence of outpatient antibiotic prescriptions associated with RSV and influenza infections in a large sample of commercially-insured children in the United States in 2008-2018.

## METHODS

### Summary: Analytic framework

In this retrospective study, we estimated the incidence and attributable fractions of antibiotic prescriptions associated with RSV and influenza. To do this, we constructed time series models regressing weekly rates of antibiotic prescriptions against RSV and influenza circulation measures estimated from this cohort and national laboratory surveillance data. We used fitted models to estimate counterfactual antibiotic prescription rates in the presence and absence of RSV and influenza circulation.

### Study population and data sources

#### Study population

Our analyses followed an open cohort of children (aged 0-17 years) captured in the Optum Clinformatics™ DataMart (“Clinformatics”) between January 1, 2008 and December 31, 2018. Clinformatics contains de-identified medical claims and enrollment data for >81 million individuals with commercial insurance annually, encompassing data on enrollee demographics and healthcare utilization including medical visits (with associated diagnoses and services) and outpatient pharmacy prescription dispenses.^19^ To mitigate immortal time bias from children with medical but not prescription capture, we defined the cohort eligible for study inclusion each week as all children with ≥1 claim for any medication within one year before or after that week. As changes due to the COVID-19 pandemic could impact this bias correction in 2019, we ended the study period in 2018. We classified age based on birth year, as birth dates are masked for privacy protection.

Analysis of these deidentified data met the definition of non-human subjects research according to the policies of the UC Berkeley Committee for the Protection of Human Subjects; Institutional Review Board review was not required.

#### Outcome definitions

Study outcomes included all-cause and cause-specific outpatient pharmacy prescription dispensing events of antibiotic agents recommended for the management of pediatric AOM, pharyngitis, sinusitis, and pneumonia (**Table S1**). We created time series including (1) all dispensed prescriptions of such antibiotics; (2) dispensed prescriptions linked to ARIs, defined as those occurring on or within 3 days of an ARI diagnosis; and (3) dispensed prescriptions linked to specific ARI syndromes including pneumonia, AOM, sinusitis, pharyngitis, non-suppurative otitis media, bronchitis, bronchiolitis, allergy, asthma, viral upper respiratory infection (URI), influenza infection, and RSV infection (**Table S2**). Prescriptions could be linked with >1 syndrome. In defining prescriptions linked to ARIs, we excluded those occurring on or within 3 days after other diagnoses for which antibiotics would likely be indicated: urinary tract, skin and soft tissue, gastrointestinal, and other bacterial infections. Additionally, in defining prescriptions linked to specific ARI syndromes for which antibiotics are not indicated (bronchiolitis, bronchitis, allergy, asthma, non-suppurative otitis media, viral URIs, RSV, influenza), we excluded those linked to ARI syndromes for which antibiotics may be indicated (pneumonia, AOM, sinusitis, pharyngitis). We defined syndromes using *International Classification of Diseases 9^th^ and 10^th^ revision, Clinical Modification* (ICD-9-CM, ICD-10-CM) codes adapted from previously-published categorizations.^15,16^

We stratified outcomes by antibiotic class (penicillin, extended-spectrum beta-lactams, macrolides, cephalosporins, tetracyclines, fluroquinolones, and other antibiotics; **Table S3**), age category (0-2, 3-5, 6-9, 10-13, and 14-17 years) and census division.

#### RSV and influenza activity

Clinical diagnosis-based and laboratory measures of RSV and influenza transmission have limitations;^20,21^ thus, we evaluated multiple measures. We considered both syndromic (medical visits with RSV/influenza diagnoses) and laboratory measures to proxy RSV and influenza circulation and selected between these based on Bayesian information criterion (BIC) values from fitted models as described below. For syndromic measures of RSV activity, we generated weekly time series of incidence of medical visits with RSV-specific diagnoses among 1) all children 0-17 years and 2) children aged 0-2 years in the study population (**Table S4**). As syndromic measures of influenza activity, we generated weekly time series of incidence of visits with influenza-specific diagnoses among 1) all children 0-17 years and 2) by age category. Age-specific measures of RSV activity were not considered for ages >2 years given limited RSV testing and use of RSV-specific diagnoses codes for this age group.^22^ As laboratory measures, we generated weekly time series of proportions of respiratory samples positive for RSV and influenza (A and B) measured from public health and clinical laboratories as reported to the National Respiratory and Enteric Virus Surveillance System (NREVSS).^23^

As RSV testing and diagnosis may vary over time, we additionally conducted sensitivity analyses using the incidence of bronchiolitis in children aged 0-2 years in the study population as the measure of RSV activity. Bronchiolitis has been considered a proxy for RSV in previous time series studies.^12,24^ We did not consider all-cause bronchiolitis codes as a measure of RSV transmission in primary analyses because, although RSV is the predominant etiology of bronchiolitis, the condition can be caused by other pathogens including human metapneumovirus, coronaviruses, and parainfluenza viruses.^25,26^ Thus, RSV-specific measures were expected to better distinguish timing of RSV and other respiratory virus circulation.

### Statistical estimation of RSV- and influenza-associated antibiotic prescriptions

We used negative binomial regression models to quantify associations between antibiotic prescriptions and measures of RSV and influenza activity. Outcome variables were weekly time series of antibiotic prescriptions. Member person-time was included as an offset term. Time series of RSV and influenza activity were included together as predictors to enable estimation of independent contributions of each viral pathogen to the outcome.^12^ Additionally, models controlled for census division, age group, time since PCV13 introduction (categorized as pre-PCV13 [2008-2009], early PCV13 period [2010–2014], and late PCV13 period [2015–2018]), as well as harmonic terms capturing seasonal patterns due to causes other than RSV and influenza (e.g., circulation of other respiratory infections). Last, we included secular trend terms to capture additional temporal trends driven by longer-term changes in the epidemiology of the conditions or in testing, coding, or prescribing practices. This yielded the general form

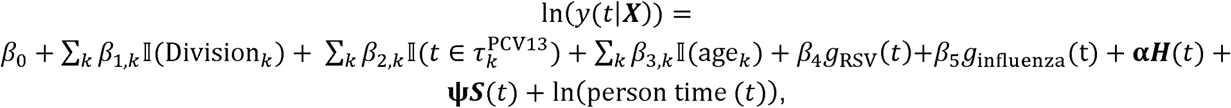

where *y* represents total weekly antibiotic prescriptions. We define *g*_RSV_(*t*) and *g*_influenza_(*t*) as measures of RSV and influenza activity at time *t,* respectively; ***H***(*t*) represents all included harmonic terms (sine and cosine); and ***S***(*t*) represents all included secular trend terms for any model. We identified the best-fitting model describing overall antibiotic prescriptions with respect to the inclusion of syndromic or laboratory-based measures of RSV and influenza activity (allowing for lag times or lead times of up to 2 weeks), harmonics (12-month, 6-month, 4-month, and 3-month), and secular trend terms (untransformed, square root-transformed, or quadratic transformation) via the BIC. We tested lag (antibiotic prescribed after RSV/influenza measures) and lead times (antibiotic prescribed before RSV/ influenza measures) to allow for variable timing in testing, presentation to care, and antibiotic prescription. The best-fitting model included the proportion of NREVSS respiratory samples positive for RSV lagged one week after antibiotic prescription as the RSV circulation measure, age-category specific influenza incidence in the study population as the influenza circulation measure, 12-month, 6-month, 4-month, and 3-month harmonics, and an untransformed secular trend term in addition to division, PCV13 period, and age category as explanatory variables. Using this best-fitting model, we fit separate models for each antibiotic class, age group, and census division, and for antibiotic prescriptions linked to any cause, any ARI diagnosis, and specific ARI syndromes.

We estimated the incidence of RSV- and influenza-associated antibiotic prescriptions as the difference between projected incidence rates from our models under scenarios with and without influenza or RSV circulation. Predicted values without circulation were estimated setting RSV or influenza terms to 0. To propagate uncertainty in attributable proportion estimates, we drew 10,000 samples from the distributions of counterfactual projected incidence rates

We conducted sensitivity analyses to verify robustness of our results. We fit linear models assuming an additive relationship between the independent and dependent variables. Additionally, we repeated primary analyses 1) including all antibiotics (not limited to respiratory agents), 2) without excluding antibiotic prescriptions linked to ARI diagnoses for which antibiotics may be indicated, and 3) broadening the sample to children who received any prescription within ±2 years of each study week.

## RESULTS

During the study period, 11,568,655 children contributed 959,237,523 person-weeks. In total, 21,365,907 outpatient antibiotic prescriptions were dispensed, yielding an incidence rate of 1,161 (95% confidence interval [CI] 1,096-1,228) prescriptions per 1,000 person-years (**Table 1; Table S5**). Two-thirds of all antibiotic prescriptions (66.8%) were linked with an ARI diagnosis; AOM, pharyngitis, and viral URI were the most common diagnoses (**Table 1**). Weekly antibiotic incidence and RSV and influenza circulation measures follow highly seasonal patterns with peaks in winter months (**Figure 1**).

**Figure 1.**
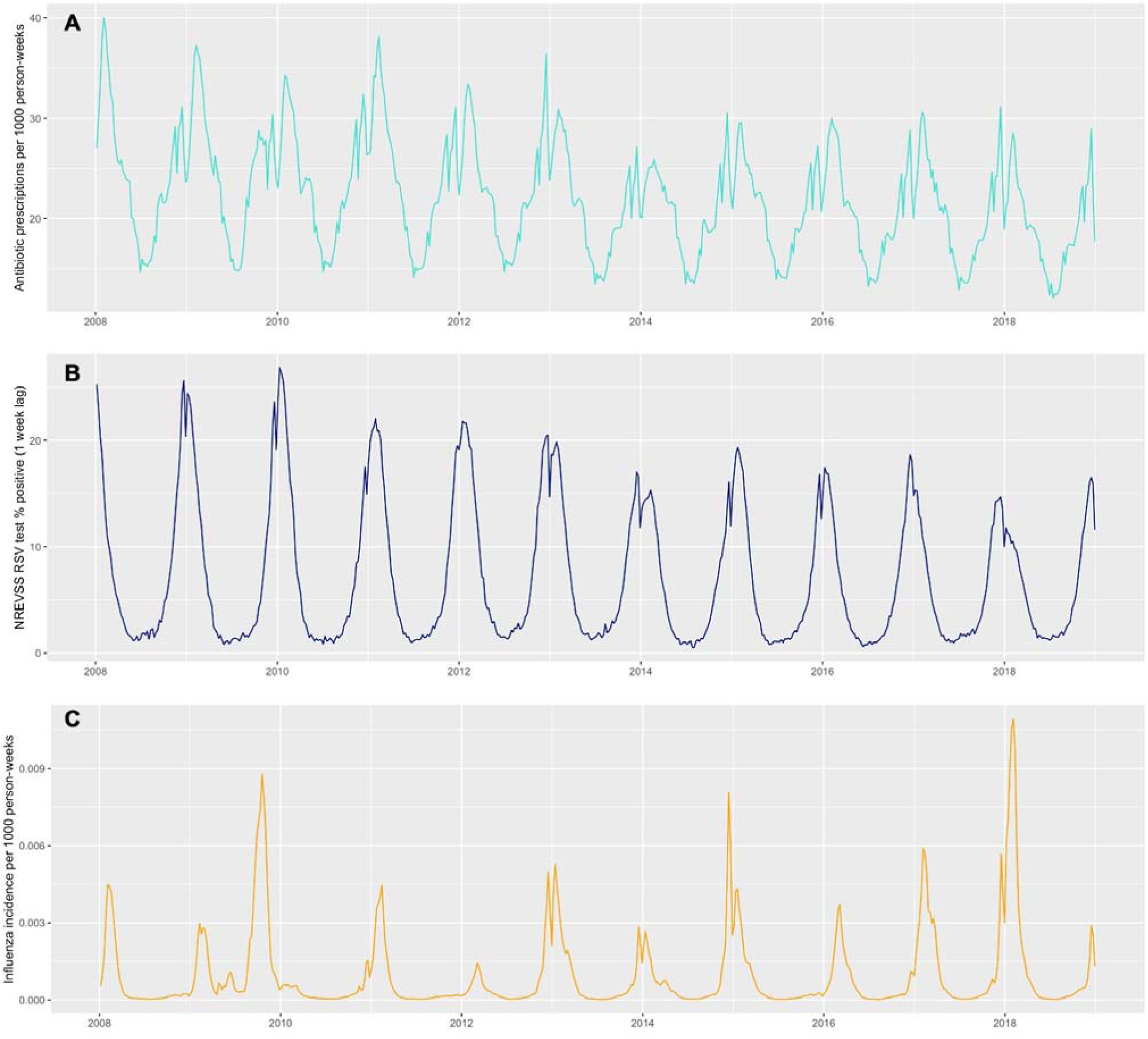
Times series of **(A)** Antibiotic prescription incidence, (**B**) RSV percent positive measure (1-week lag), and (**C**) influenza visit incidence (all ages)

**Table 1:**
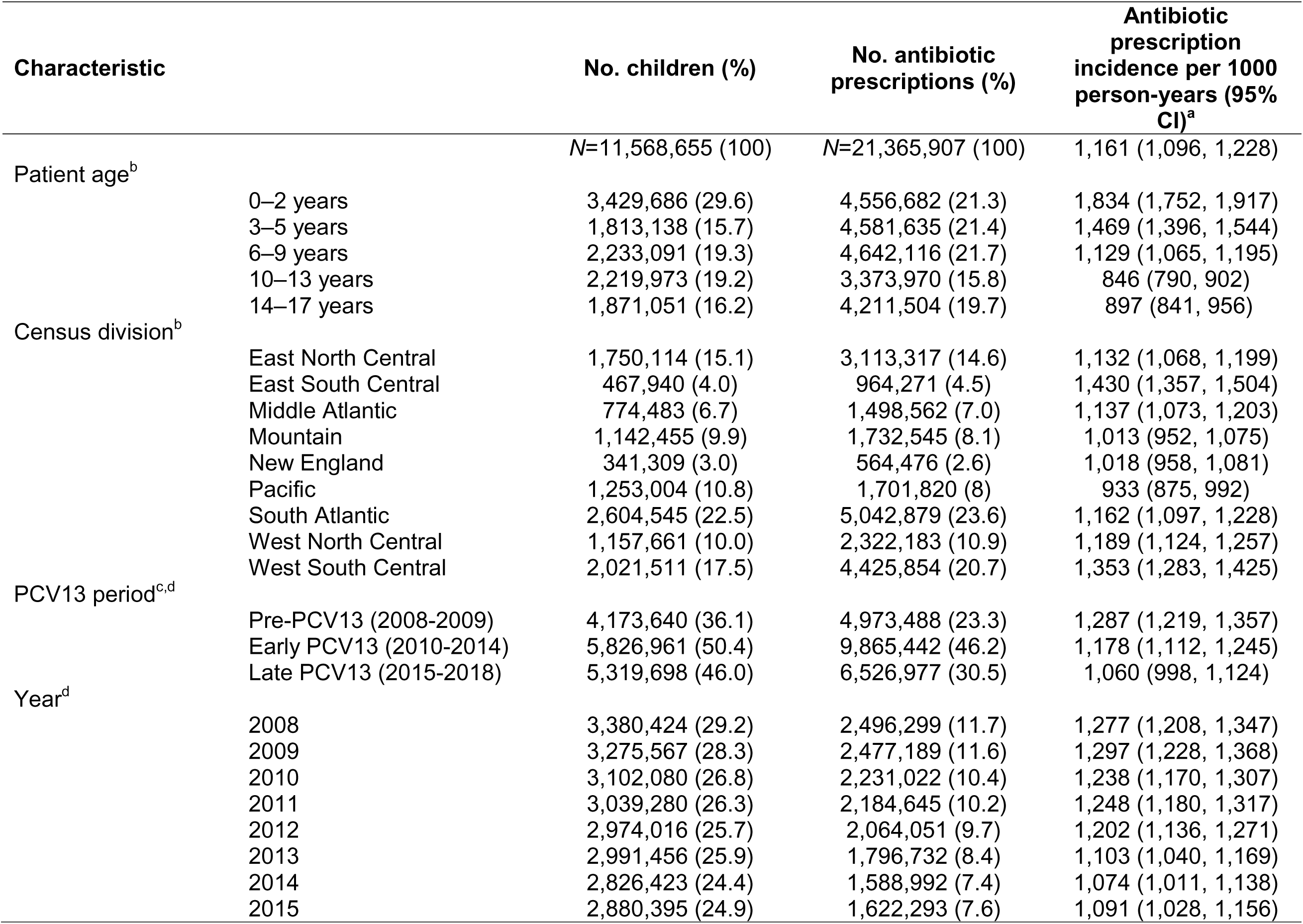
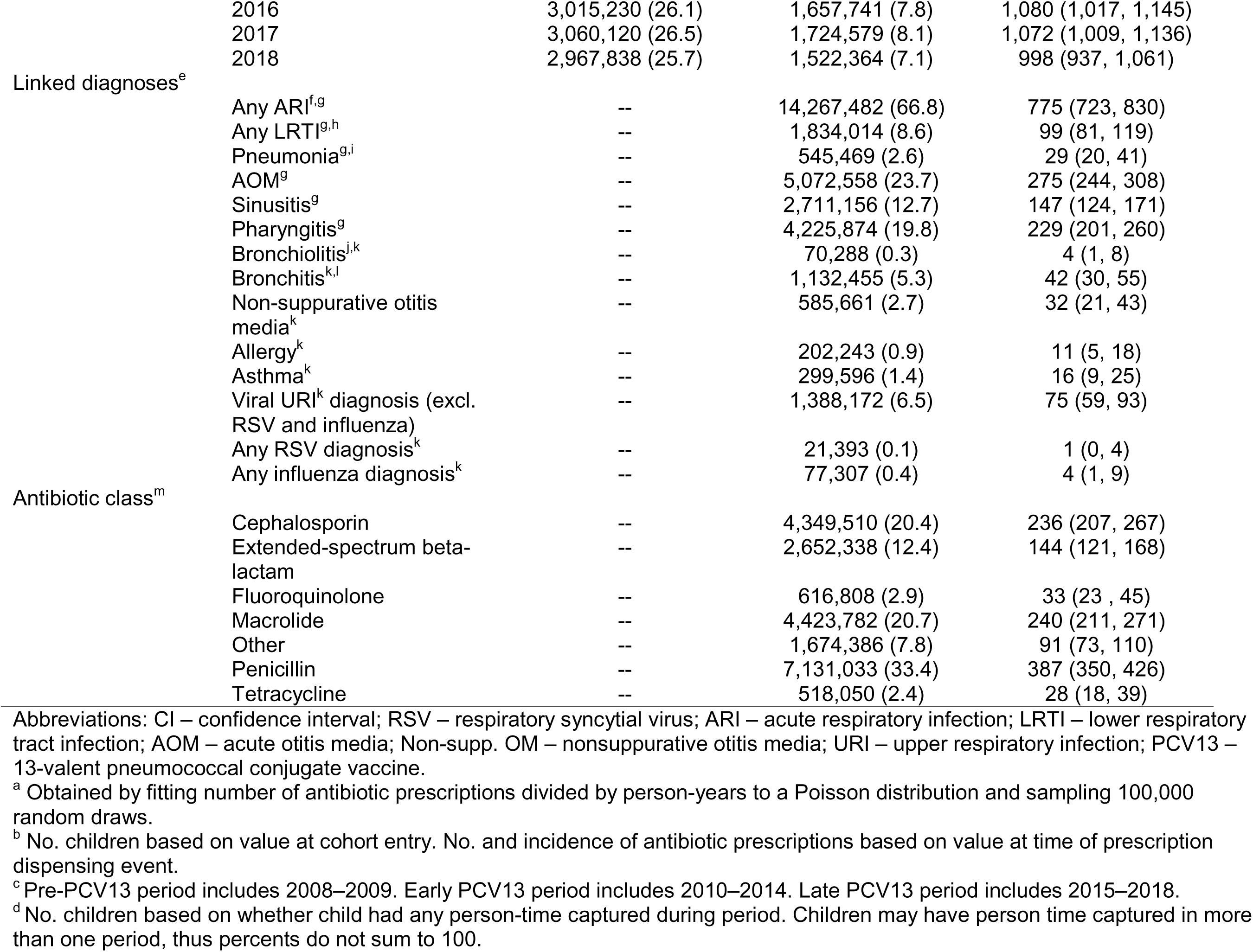
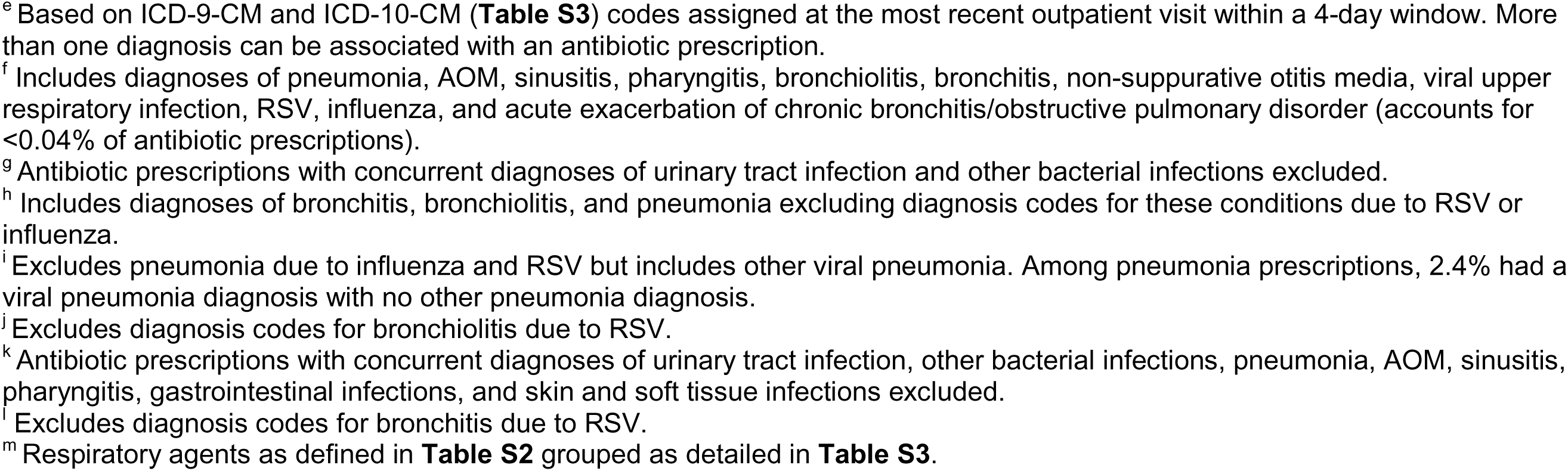
Cohort and outpatient antibiotic prescriptions by prescription and patient characteristics, Optum Clinformatics 2008-2018.

We estimated that 6.3% (5.2-7.3%) of all antibiotic prescriptions were associated with RSV (**Table 2**). This fraction varied by week and exceeded 10% during periods of peak RSV circulation each year (**Figure 2**). RSV-associated proportions were highest among young children, with 8.6% (5.6-11.4%) and 8.4% (5.6-11.3%) of antibiotic prescriptions associated with RSV among children aged 0-2 years and 3-5 years, respectively. Among antibiotics linked to ARI and LRTI diagnoses, 8.1% (6.7-9.5%) and 12.5% (10.8-14.2%) of prescriptions were associated with RSV. Aside from RSV-specific diagnoses, the conditions for which RSV accounted for the greatest proportion of antibiotic prescriptions included bronchiolitis (19.3% [16.8-21.7]%), bronchitis (9.6% [7.5-11.7%]), and pneumonia (11.7% [10.0-13.3%]). In sensitivity analyses, models measuring RSV activity using bronchiolitis visits yielded higher RSV-associated proportion estimates compared with estimates from primary analyses, with 8.0% (6.8-9.3%) of antibiotic prescriptions in children associated with RSV (**Table S6**).

**Figure 2.**
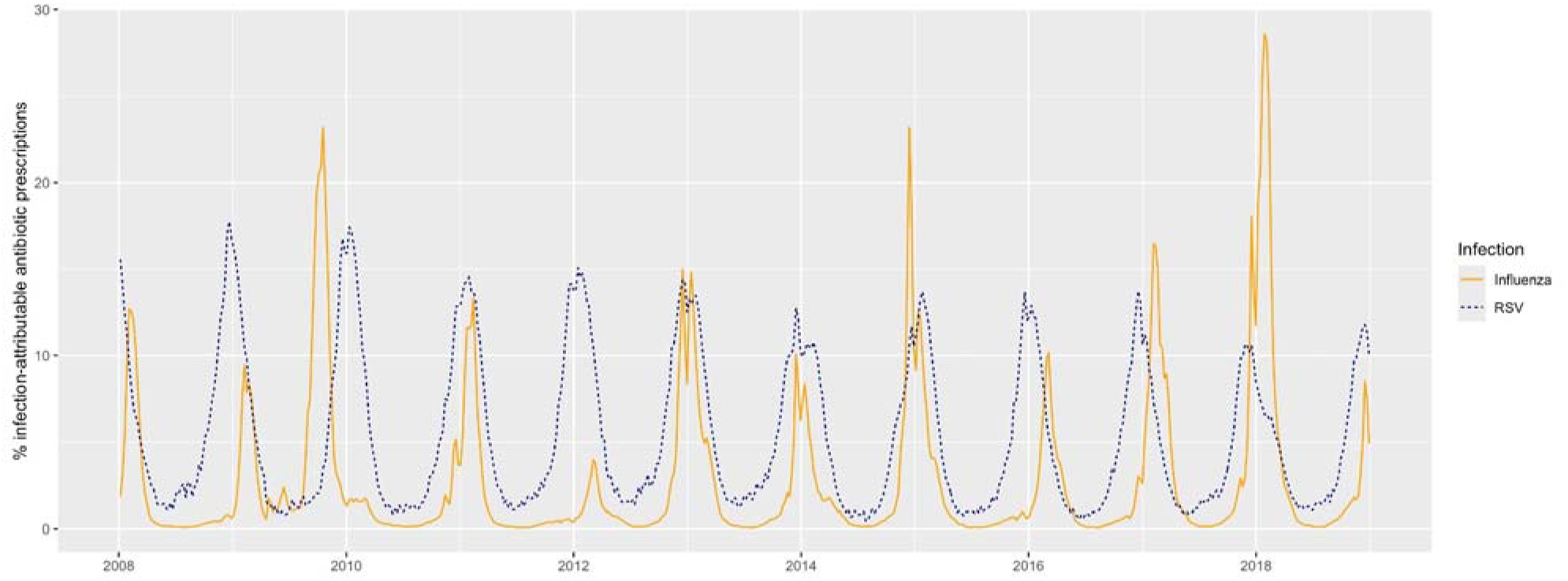
Proportion of antibiotic prescriptions associated with RSV and influenza by week

**Table 2:**
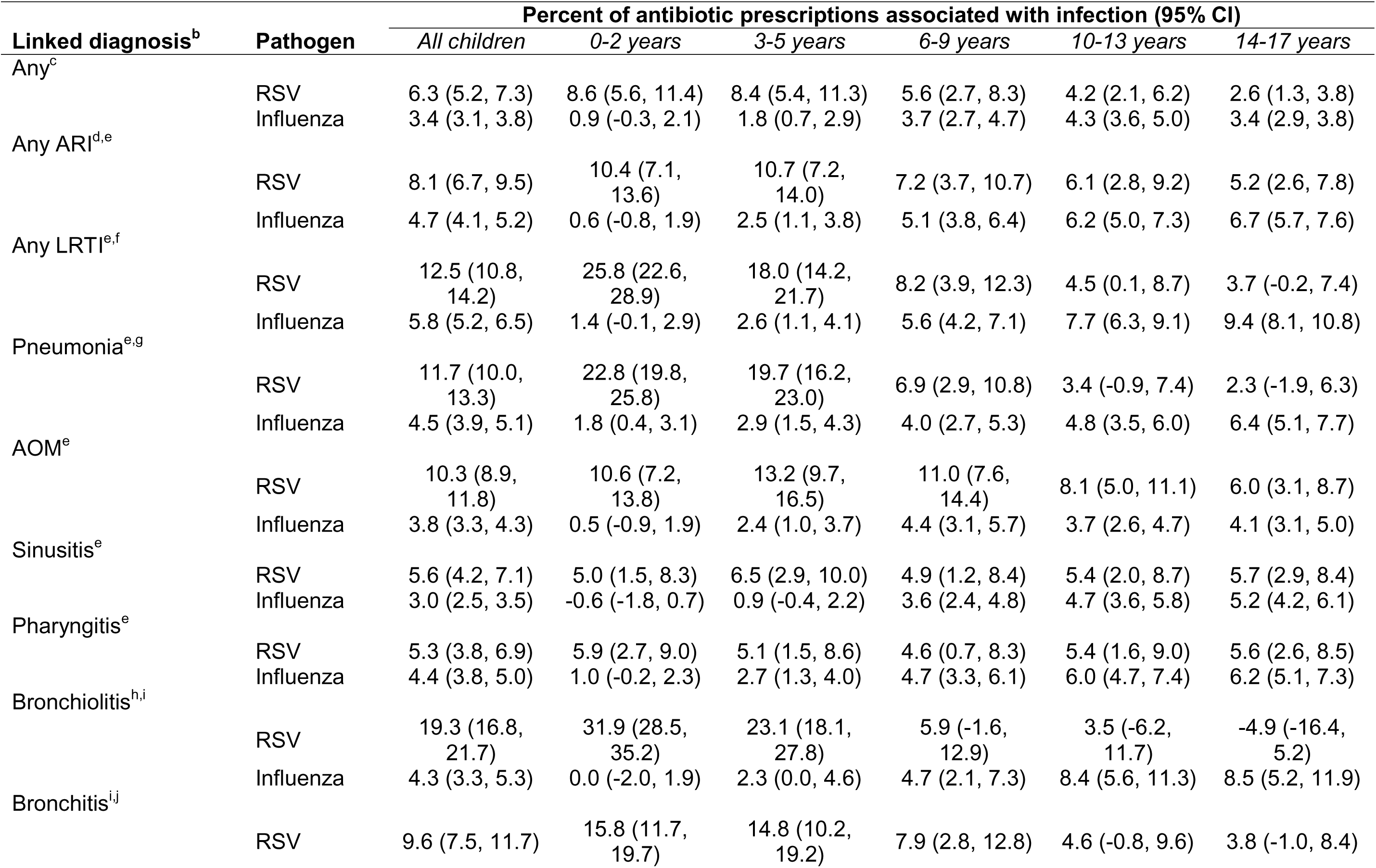
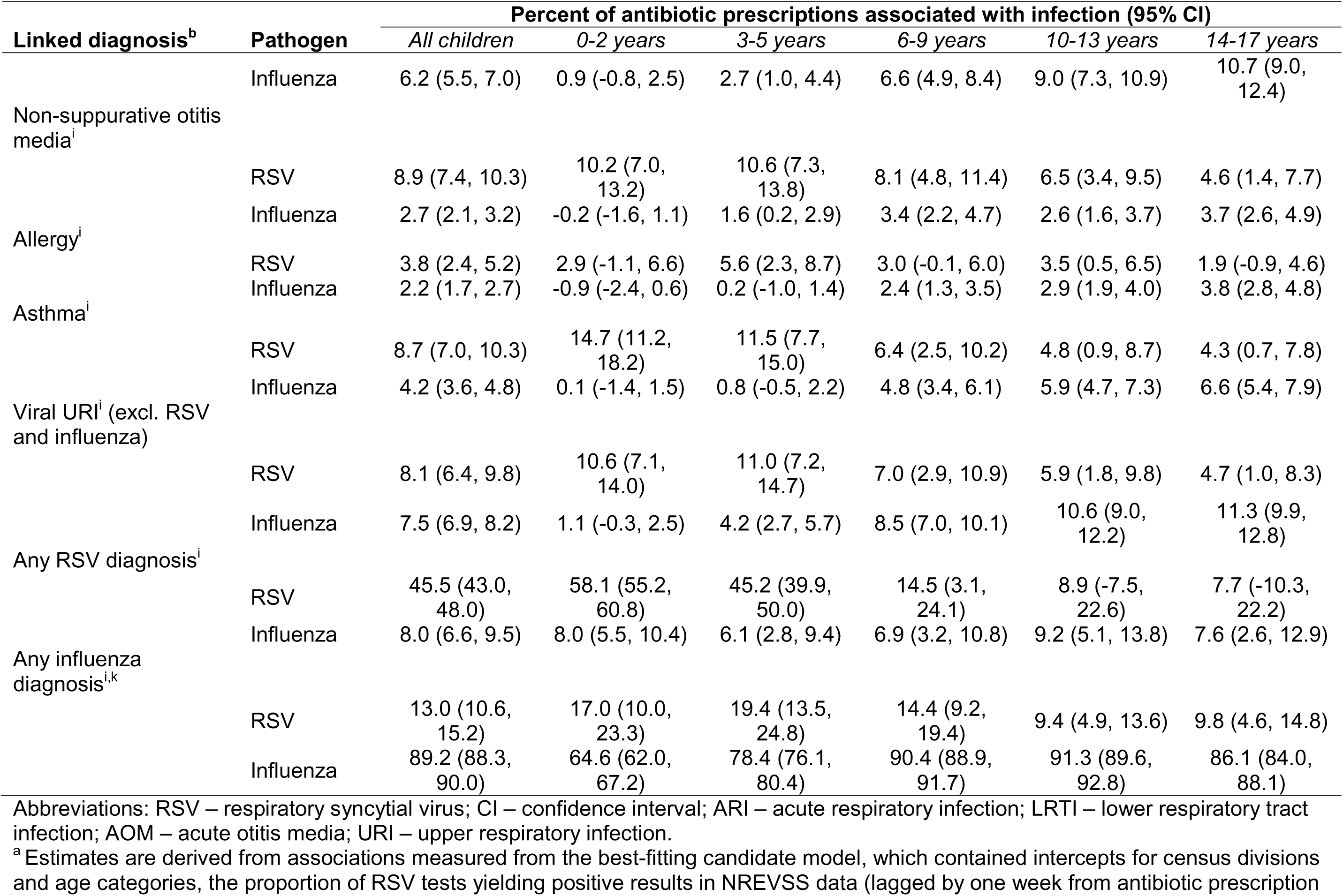
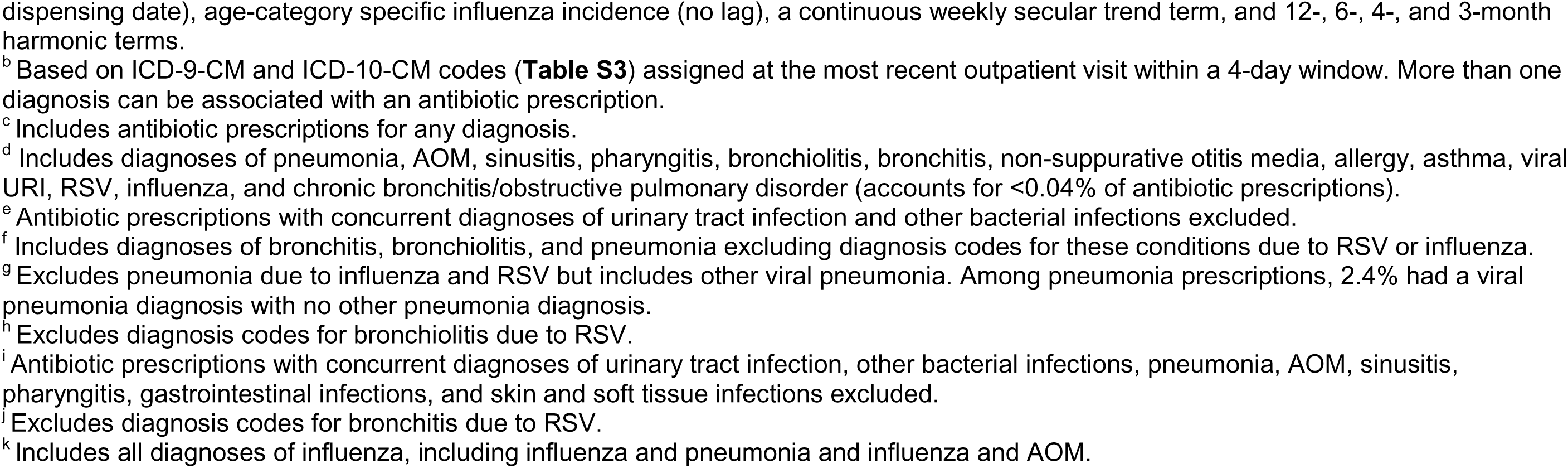
Model-estimated^a^ proportion of outpatient antibiotic prescriptions associated with respiratory syncytial virus infection, by age group and linked diagnoses among children 0-17 years, 2008-2018.

Incidence rates of RSV-associated antibiotic prescriptions among children were 72.6 (59.7-85.9) and 62.7 (51.2-74.6) per 1000 person-years for all causes and for ARIs, respectively (**Table 3**). Children aged 0-2 years experienced the highest incidence of RSV-associated antibiotic prescriptions, with 157.1 (102.6-211.2) prescriptions per 1000 person-years. By syndrome, the highest incidence of RSV-associated antibiotic prescriptions was linked with AOM diagnoses (28.4 [23.4, 34.0] per 1000 person-years). We estimated that 49.3 (39.7-59.6) RSV-associated antibiotic prescriptions per 1000 person-years were linked to ≥1 ARI diagnosis for which antibiotics may be indicated, while 12.6 (9.8-15.9) RSV-associated antibiotic prescriptions per 1000 person-years were linked only with ARI diagnoses for which antibiotics are not guideline-recommended.

**Table 3:**
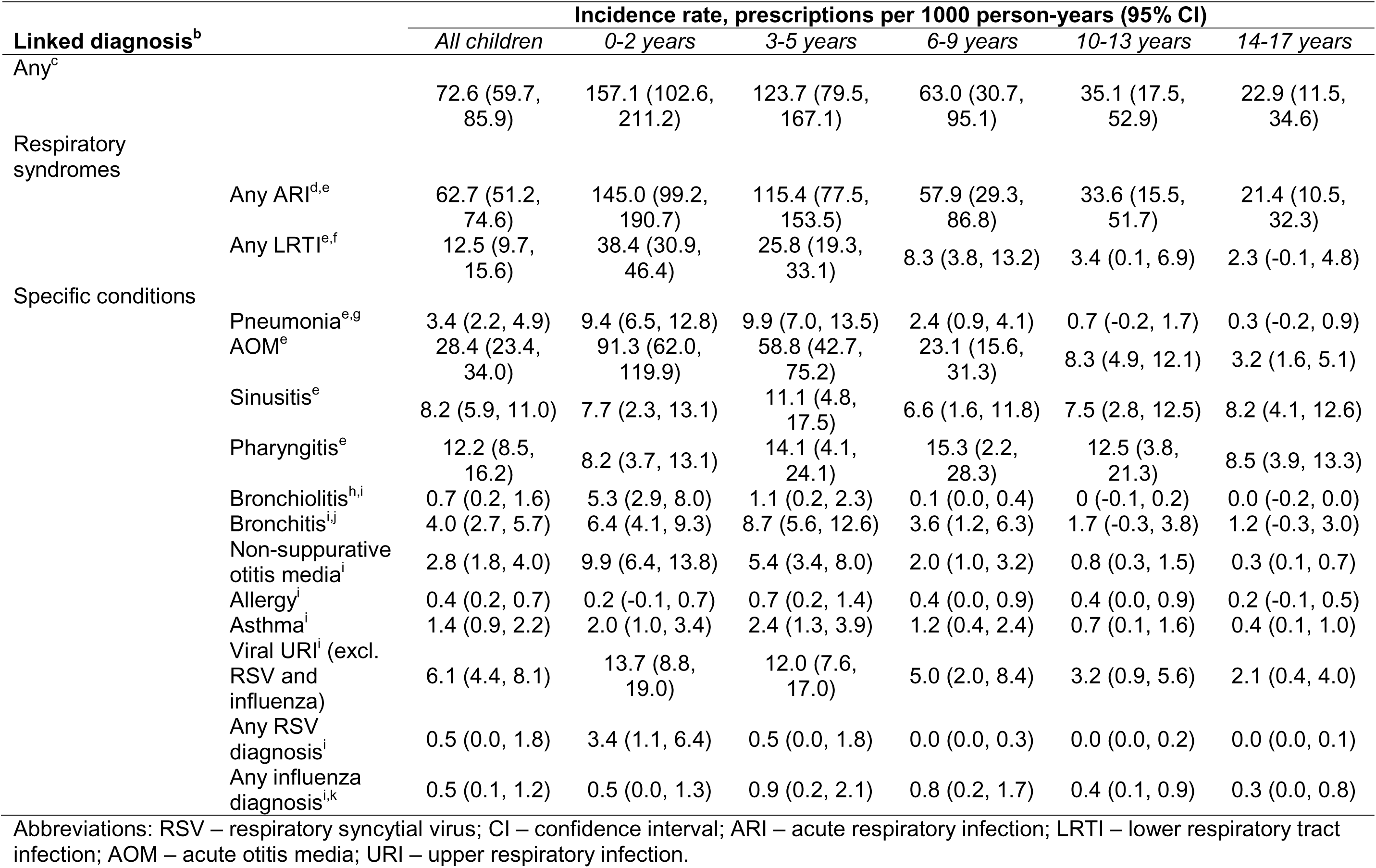
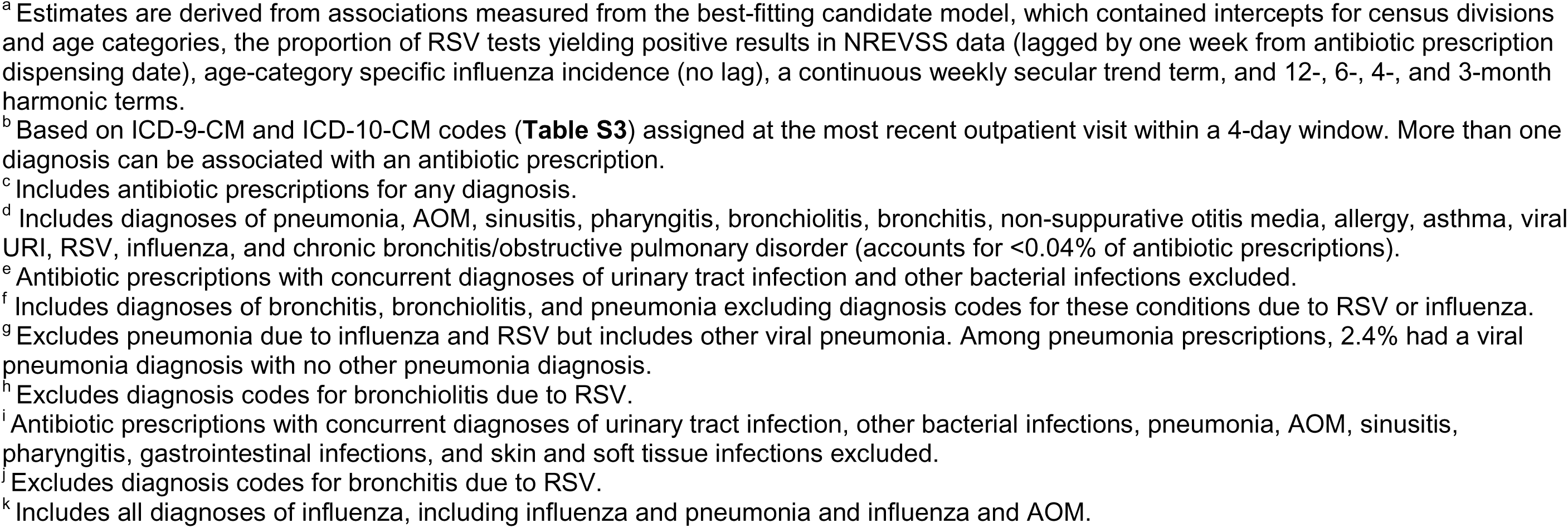
Model-estimated^a^ incidence of antibiotic prescriptions associated with respiratory syncytial virus infection by age group and diagnosis among children 0-17 years, 2008-2018.

| Linked diagnosis<sup>b</sup> |  | Incidence rate, prescriptions per 1000 person-years (95% CI) |  |  |  |  |  |
| --- | --- | --- | --- | --- | --- | --- | --- |
|  |  | *All children* | *0-2 years* | *3-5 years* | *6-9 years* | *10-13 years* | *14-17 years* |
| Any<sup>c</sup> |  | 72.6 (59.7, 85.9) | 157.1 (102.6, 211.2) | 123.7 (79.5, 167.1) | 63.0 (30.7, 95.1) | 35.1 (17.5, 52.9) | 22.9 (11.5, 34.6) |
| Respiratory syndromes |  |  |  |  |  |  |  |
|  | Any ARI<sup>d,e</sup> | 62.7 (51.2, 74.6) | 145.0 (99.2, 190.7) | 115.4 (77.5, 153.5) | 57.9 (29.3, 86.8) | 33.6 (15.5, 51.7) | 21.4 (10.5, 32.3) |
|  | Any LRTI<sup>e,f</sup> | 12.5 (9.7, 15.6) | 38.4 (30.9, 46.4) | 25.8 (19.3, 33.1) | 8.3 (3.8, 13.2) | 3.4 (0.1, 6.9) | 2.3 (-0.1, 4.8) |
| Specific conditions |  |  |  |  |  |  |  |
|  | Pneumonia<sup>e,g</sup> | 3.4 (2.2, 4.9) | 9.4 (6.5, 12.8) | 9.9 (7.0, 13.5) | 2.4 (0.9, 4.1) | 0.7 (-0.2, 1.7) | 0.3 (-0.2, 0.9) |
|  | AOM<sup>e</sup> | 28.4 (23.4, 34.0) | 91.3 (62.0, 119.9) | 58.8 (42.7, 75.2) | 23.1 (15.6, 31.3) | 8.3 (4.9, 12.1) | 3.2 (1.6, 5.1) |
|  | Sinusitis<sup>e</sup> | 8.2 (5.9, 11.0) | 7.7 (2.3, 13.1) | 11.1 (4.8, 17.5) | 6.6 (1.6, 11.8) | 7.5 (2.8, 12.5) | 8.2 (4.1, 12.6) |
|  | Pharyngitis<sup>e</sup> | 12.2 (8.5, 16.2) | 8.2 (3.7, 13.1) | 14.1 (4.1, 24.1) | 15.3 (2.2, 28.3) | 12.5 (3.8, 21.3) | 8.5 (3.9, 13.3) |
|  | Bronchiolitis<sup>h,i</sup> | 0.7 (0.2, 1.6) | 5.3 (2.9, 8.0) | 1.1 (0.2, 2.3) | 0.1 (0.0, 0.4) | 0 (-0.1, 0.2) | 0.0 (-0.2, 0.0) |
|  | Bronchitis<sup>i,j</sup> | 4.0 (2.7, 5.7) | 6.4 (4.1, 9.3) | 8.7 (5.6, 12.6) | 3.6 (1.2, 6.3) | 1.7 (-0.3, 3.8) | 1.2 (-0.3, 3.0) |
|  | Non-suppurative otitis media<sup>i</sup> | 2.8 (1.8, 4.0) | 9.9 (6.4, 13.8) | 5.4 (3.4, 8.0) | 2.0 (1.0, 3.2) | 0.8 (0.3, 1.5) | 0.3 (0.1, 0.7) |
|  | Allergy<sup>i</sup> | 0.4 (0.2, 0.7) | 0.2 (-0.1, 0.7) | 0.7 (0.2, 1.4) | 0.4 (0.0, 0.9) | 0.4 (0.0, 0.9) | 0.2 (-0.1, 0.5) |
|  | Asthma<sup>i</sup> | 1.4 (0.9, 2.2) | 2.0 (1.0, 3.4) | 2.4 (1.3, 3.9) | 1.2 (0.4, 2.4) | 0.7 (0.1, 1.6) | 0.4 (0.1, 1.0) |
|  | Viral URI<sup>i</sup> (excl. RSV and influenza) | 6.1 (4.4, 8.1) | 13.7 (8.8, 19.0) | 12.0 (7.6, 17.0) | 5.0 (2.0, 8.4) | 3.2 (0.9, 5.6) | 2.1 (0.4, 4.0) |
|  | Any RSV diagnosis<sup>i</sup> | 0.5 (0.0, 1.8) | 3.4 (1.1, 6.4) | 0.5 (0.0, 1.8) | 0.0 (0.0, 0.3) | 0.0 (0.0, 0.2) | 0.0 (0.0, 0.1) |
|  | Any influenza diagnosis<sup>l,k</sup> | 0.5 (0.1, 1.2) | 0.5 (0.0, 1.3) | 0.9 (0.2, 2.1) | 0.8 (0.2, 1.7) | 0.4 (0.1, 0.9) | 0.3 (0.0, 0.8) |

Abbreviations: RSV – respiratory syncytial virus; CI – confidence interval; ARI – acute respiratory infection; LRTI – lower respiratory tract infection; AOM – acute otitis media; URI – upper respiratory infection.
<sup>a</sup> Estimates are derived from associations measured from the best-fitting candidate model, which contained intercepts for census divisions and age categories, the proportion of RSV tests yielding positive results in NREVSS data (lagged by one week from antibiotic prescription dispensing date), age-category specific influenza incidence (no lag), a continuous weekly secular trend term, and 12-, 6-, 4-, and 3-month harmonic terms.
<sup>b</sup> Based on ICD-9-CM and ICD-10-CM codes (**Table S3**) assigned at the most recent outpatient visit within a 4-day window. More than one diagnosis can be associated with an antibiotic prescription.
<sup>c</sup> Includes antibiotic prescriptions for any diagnosis.
<sup>d</sup> Includes diagnoses of pneumonia, AOM, sinusitis, pharyngitis, bronchiolitis, bronchitis, non-suppurative otitis media, allergy, asthma, viral URI, RSV, influenza, and chronic bronchitis/obstructive pulmonary disorder (accounts for <0.04% of antibiotic prescriptions).
<sup>e</sup> Antibiotic prescriptions with concurrent diagnoses of urinary tract infection and other bacterial infections excluded.
<sup>f</sup> Includes diagnoses of bronchitis, bronchiolitis, and pneumonia excluding diagnosis codes for these conditions due to RSV or influenza.
<sup>g</sup> Excludes pneumonia due to influenza and RSV but includes other viral pneumonia. Among pneumonia prescriptions, 2.4% had a viral pneumonia diagnosis with no other pneumonia diagnosis.
<sup>h</sup> Excludes diagnosis codes for bronchiolitis due to RSV.
<sup>i</sup> Antibiotic prescriptions with concurrent diagnoses of urinary tract infection, other bacterial infections, pneumonia, AOM, sinusitis, pharyngitis, gastrointestinal infections, and skin and soft tissue infections excluded.
<sup>j</sup> Excludes diagnosis codes for bronchitis due to RSV.
<sup>k</sup> Includes all diagnoses of influenza, including influenza and pneumonia and influenza and AOM.

Overall, we estimated that 3.4% (3.1-3.8%) of all antibiotic prescriptions and 4.7% (4.1-5.2%) of ARI-associated prescriptions were associated with influenza (**Table 2**). This estimate varied considerably across years; during periods of peak circulation, influenza-associated prescriptions accounted for >20% of antibiotic prescribing in 2009-10, 2015-16, and 2017-18, but <10% of prescribing during seasonal peaks in 2008-09 and 2011-12 (**Figure 2**). Influenza accounted for a greater proportion of antibiotic prescriptions among children aged ≥6 years than among those aged 0-5 years. Aside from influenza-specific diagnoses, diagnoses for which influenza accounted for the greatest proportions of antibiotic prescriptions included bronchitis (6.2% [5.5-7.0%]), viral URI (7.5% [6.9-8.2%]), and physician-coded RSV (8.1% [6.6-9.5%]). Estimates of influenza-associated antibiotic prescriptions changed minimally in sensitivity analyses using bronchiolitis to proxy RSV (**Table S7**).

Influenza-associated antibiotic prescription incidence rates were 40.0 (35.1-45.1) and 36.2 (31.6-41.1) per 1000 person-years for all causes and for ARIs, respectively (**Table 4**). Rates were highest for prescriptions associated with AOM (10.5 [8.6-12.5]) and pharyngitis (10.2 [8.4-12.2]) and were greater at ages ≥6 years compared with 0-5 years. We estimated that 25.8 (21.8-29.9) influenza-associated antibiotic prescriptions per 1000 person-years were linked to ≥1 ARI diagnosis for which antibiotics may be indicated, while 11.2 (9.3-13.3) influenza-associated antibiotic prescriptions per 1000 person-years were linked only with ARI diagnoses for which antibiotics are not guideline-recommended.

**Table 4:**
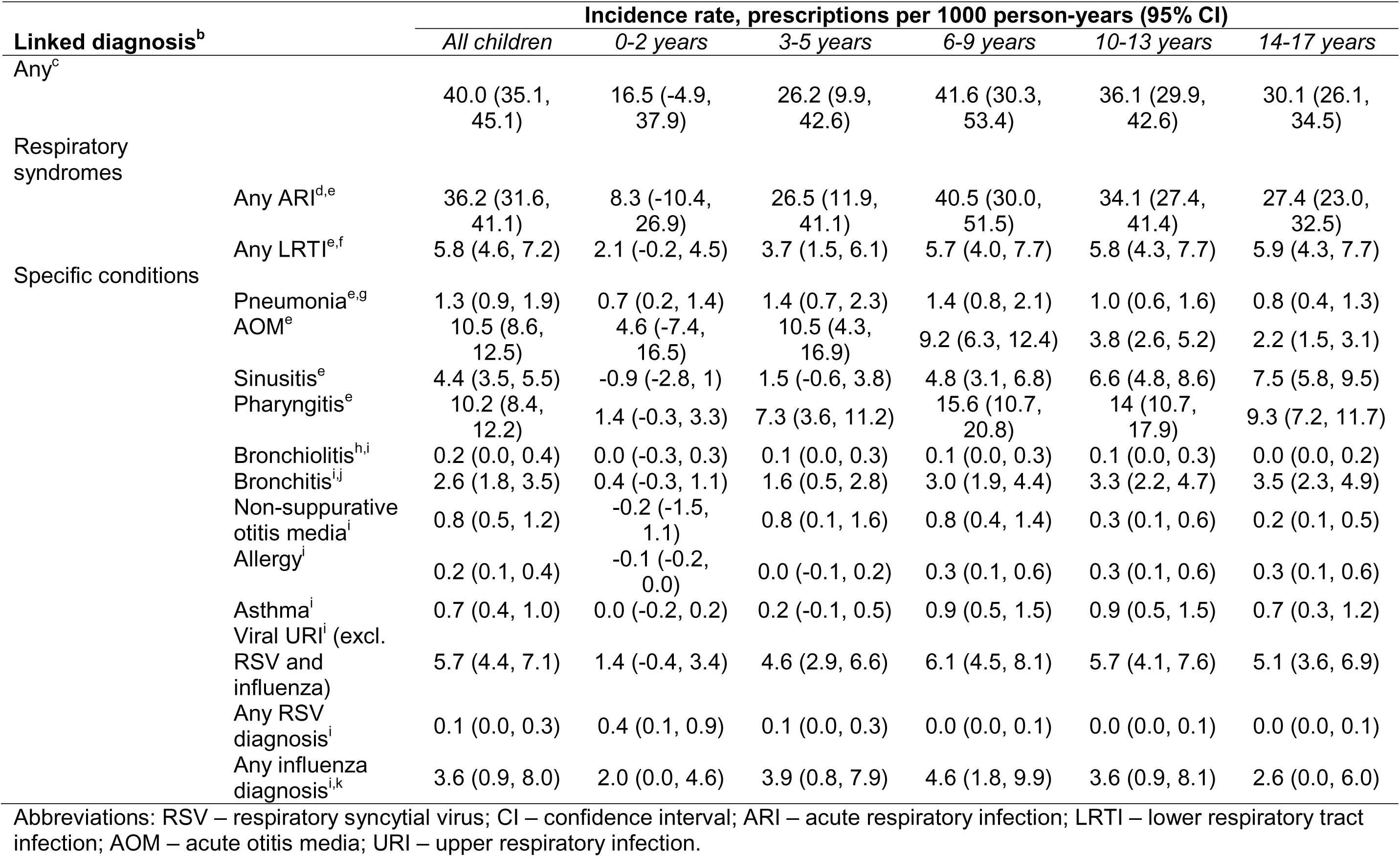
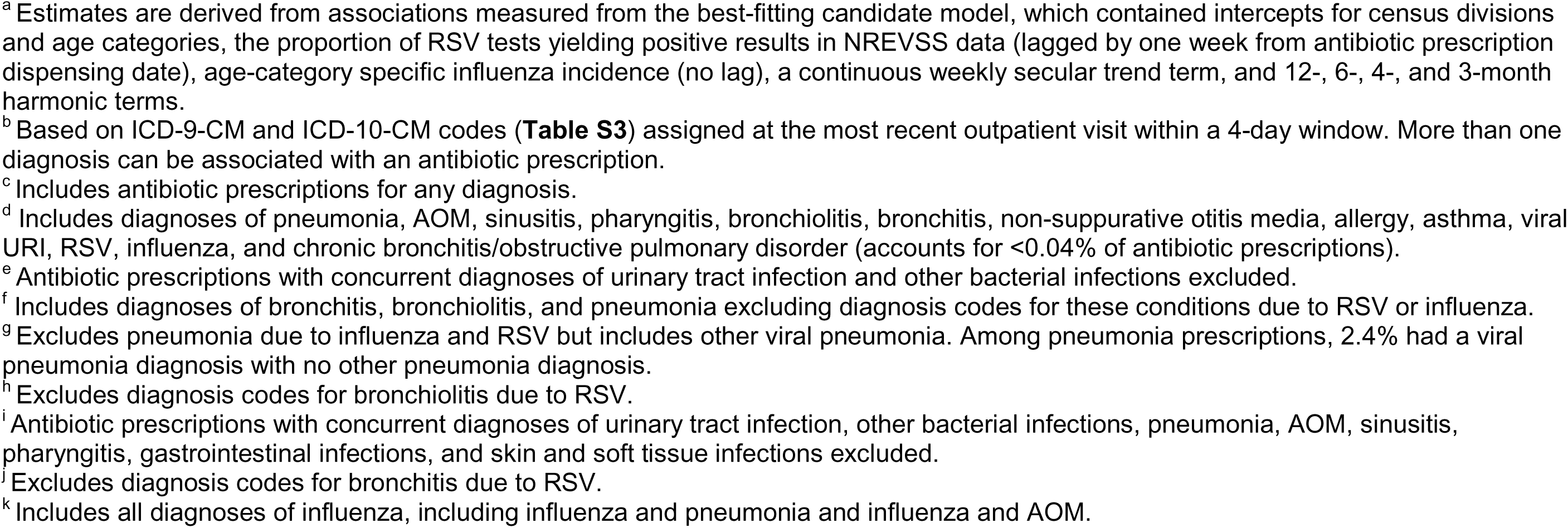
Model-estimated^a^ incidence of antibiotic prescriptions associated with influenza virus infection by age group and diagnosis among children 0-17 years, 2008-2018.

By class, the most frequently dispensed antibiotics were penicillins (33.4% of antibiotic prescriptions), followed by macrolides (20.7%), and cephalosporins (20.4%; **Table 1**). RSV was associated with >6% of penicillin, macrolide, and extended-spectrum beta-lactam prescriptions (**Table 5**). Influenza accounted for 4.4% (4.2-4.6%) of macrolide prescriptions and 3.8% (3.6-4.0%) of penicillin prescriptions.

**Table 5:** Model-estimated^a^ proportion of outpatient antibiotic prescriptions associated with respiratory syncytial virus and influenza virus infection, by antibiotic class.

| Antibiotic class<sup>a</sup> | Percent of antibiotics associated with infection (95% CI) |  | Antibiotic prescription incidence per 1000 person-years (95% CI) |  |
| --- | --- | --- | --- | --- |
|  | *RSV* | *Influenza* | *RSV* | *Influenza* |
| Macrolide | 7.9 (7.4, 8.5) | 4.4 (4.2, 4.6) | 19.1 (16.4, 22.0) | 10.7 (9.3, 12.1) |
| Extended-spectrum beta-lactam | 6.7 (6.2, 7.3) | 3.0 (2.8, 3.2) | 9.7 (8.0, 11.6) | 4.2 (3.5, 5.0) |
| Penicillin | 6.5 (6.0, 7.0) | 3.8 (3.6, 4.0) | 25.1 (22.2, 28.4) | 14.7 (13.1, 16.3) |
| Cephalosporin | 5.6 (5.1, 6.1) | 3.3 (3.1, 3.4) | 13.2 (11.3, 15.4) | 7.7 (6.7, 8.8) |
| Tetracycline | 1.5 (0.7, 2.3) | -0.1 (-0.3, 0.1) | 0.4 (0.2, 0.7) | 0.0 (-0.1, 0.0) |
| Fluoroquinolone | 0.9 (0.0, 1.8) | 0.9 (0.6, 1.2) | 0.3 (0.0, 0.6) | 0.3 (0.2, 0.5) |
| Other | 0.7 (0.1, 1.2) | 0.2 (0.0, 0.3) | 0.6 (0.1, 1.2) | 0.2 (0.0, 0.3) |

Abbreviations: CI – confidence interval; RSV – respiratory syncytial virus.
<sup>a</sup> Estimates are derived from associations measured from the best-fitting candidate model, which contained intercepts for census divisions and age categories, the proportion of RSV tests yielding positive results in NREVSS data (lagged by one week from antibiotic prescription dispensing date), age-category specific influenza incidence (no lag), a continuous weekly secular trend term, and 12-, 6-, 4-, and 3-month harmonic terms.
<sup>b</sup> Respiratory agents as defined in **Table S1** for any diagnosis as categorized in **Table S2**.

RSV and influenza visit incidence were highest in the East South Central Division (**Table S8**). Overall (**Table 1**) and RSV- and influenza-attributable antibiotic prescriptions (**Table S9**) were concentrated among divisions in the South. There was wide variation in point estimates of the proportion of antibiotic prescriptions associated with RSV, although confidence intervals overlapped. Differences in influenza-associated antibiotic prescriptions were less apparent.

Sensitivity analyses using a linear model, modifying the antibiotic definition to include all agents, and including children with pharmacy dispenses within ±2 years of each study week yielded results similar to those derived from primary analyses (**Tables S10-S12).** Including antibiotic prescriptions linked to diagnoses for which antibiotics may be indicated yielded only slight changes in the incidence of RSV-associated and influenza-associated antibiotic prescriptions linked to bronchiolitis, bronchitis, non-suppurative otitis media, allergy, asthma, and viral URI diagnoses (**Table S13**).

## DISCUSSION

We estimated that approximately 6% and 3% of antibiotic prescriptions were associated with RSV and influenza infections, respectively, among a large sample of US children from 2008-2018. These estimates translate to 73 RSV-associated antibiotic prescriptions and 40 influenza-associated antibiotic prescriptions per 1000 children annually. Most RSV- and influenza-associated prescriptions were linked with ≥1 ARI diagnosis. The incidence of RSV-associated antibiotic prescriptions was highest among children aged :::5 years and RSV accounted for large shares of antibiotic prescriptions linked to pneumonia, bronchiolitis, and AOM diagnoses. In contrast, influenza-associated antibiotic prescription rates were highest among children aged >5 years and more commonly linked with viral URI and bronchitis diagnoses.

Compared with a study of Scottish children aged <5 years, which found RSV and influenza were associated with 7% and 2%, respectively, of all antibiotic prescriptions among children aged <5 years,^12^ we estimated higher proportions of RSV-associated antibiotic prescriptions and lower proportions of influenza-associated antibiotic prescriptions at ages <5 years. A time-series analysis of antibiotic prescriptions from general practitioners in the United Kingdom found RSV accounted for 11%, 10%, 8%, and 4% of prescribing in children 0-5 months, 6-23 months, 2-4 years, and 5-14 years, respectively,^14^ aligned with estimates from our study. Compared with a study of children in Kaiser Permanente Northern California,^13^ we estimated higher proportions of influenza-associated antibiotic prescriptions. However, in our study, the Pacific division (including California) was among those with the lowest proportions of influenza-associated prescribing. Although comparison with existing studies contextualizes our findings, direct comparison is limited given geographic differences in prescribing.^15–17^

Our findings suggest that improved RSV and influenza control may reduce pediatric antibiotic use. Influenza vaccines are recommended for all children without contraindications.^27^ Immunization against RSV, via maternal vaccination (RSV preF protein vaccine) or the monoclonal antibody nirsevimab, has recently been implemented to protect against RSV among infants aged <6 months (RSV preF, nirsevimab) and children aged <20 months at risk of severe RSV disease (nirsevimab).^28,29^ Substantial vaccine-preventable burdens persist from suboptimal immunization uptake. In 2023-2024, only 56% of infants were immunized against RSV (maternal RSV preF vaccine or nirsevimab).^30^ Similarly, only 55% of children received influenza vaccine in 2023-24, with uptake lower among older children than younger children.^31^

While the pediatric age range targeted by current RSV immunization recommendations is narrow, prevention of early-life RSV infections may mitigate antibiotic use in later infancy and childhood. Studies across settings^32–34^ have demonstrated that early-life RSV infections are associated with increased risk of recurrent wheeze, asthma, and ARIs in later infancy and childhood, conditions which may precipitate antibiotic use. In a US cohort, heightened risk of conditions including AOM and pneumonia after early-life RSV infections led to a 23% overall increase in children’s odds of antibiotic treatment at ages 7-12 months.^34^ Prevention of early-life related antibiotic exposures is also independently important. Antibiotic exposures during infancy may enhance subsequent risk of chronic disease outcomes including asthma, allergic rhinitis, atopic dermatitis, and obesity.^35^

Prevention of severe early-life RSV infections is the focus of current infant immunization programs. However, RSV-attributable ARIs remain important causes of disease among older infants, toddlers, and young children. We found that RSV-associated antibiotic prescription incidence was similar at ages 0-2 and 3-5 years with 157 and 124 prescriptions per 1000 child-years, respectively. RSV immunization programs targeting children aged >12 months are in clinical trials^36,37^ and may offer strategies for preventing RSV-related antibiotic use among older children. While severe RSV-associated LRTIs are most prominent in infants and young children,^38^ numerous studies have also demonstrated that older children also acquire RSV infection and experience mild or nonspecific symptoms associated with these infections.^39–41^ Our study and earlier findings^13,14^ that RSV and influenza make contribute to antibiotic use in later childhood demonstrate the potential value of interventions that reduce virus circulation within the general population.

Prevention of RSV and influenza infections through immunizations is an important adjunct to traditional antibiotic stewardship interventions. In the US, AOM is a leading cause of pediatric antibiotic use^3,15,16^ where almost one-third of AOM-associated prescriptions are guideline non-concordant.^42^ In our study, RSV and influenza accounted for 10% and 4% of antibiotic prescriptions linked with AOM, respectively. Additionally, antibiotic treatment of conditions for which antibiotics are not indicated is common, although decreases have been noted.^3,15,16^ We estimated that RSV and influenza accounted for approximately 13 and 11 antibiotic prescriptions per 1000 person-years, respectively, for which antibiotics are not guideline-recommended. Stewardship interventions targeting ARI-associated prescribing have demonstrated improvements in guideline-concordant care,^43^ but require ongoing investment. We also found that macrolides were the antibiotic class for which RSV and influenza accounted for the greatest shares of all prescriptions (8% and 4%, respectively). Reduction of such prescribing is important, as macrolides are frequently prescribed to children for ARIs for which antibiotics are not recommended.^16^

This study has limitations. First, we considered broad age classes. Differences in RSV infection risk and clinical outcomes by month of life during infancy may be relevant to anticipating impacts of maternal vaccination and nirsevimab on antibiotic use.^11^ The previously-discussed UK study found the highest proportions of RSV-attributable antibiotic prescriptions among children 0-5 months,^14^ suggesting potential reductions in RSV-associated antibiotics from infant immunization in this age group may be even greater than those expected based on our findings. Second, we used measures of virus activity aggregated across ages within census divisions, which may mask variation at smaller scales^44^ and across age groups.^45^ Third, we relied on ICD-9-CM and ICD-10-CM codes to assign diagnoses to antibiotic prescriptions. Providers may be incentivized to assign antibiotic-appropriate codes to visits in which antibiotics are prescribed, which could affect evaluation of antibiotics by diagnosis. However, potential code-shifting would likely not have affected estimates of overall and all ARI-linked antibiotic prescriptions. Fourth, we included data from before the COVID-19 pandemic to capture typical seasonal endemic patterns of RSV and influenza activity. Pandemic-related pathogen circulation disruptions^46,47^ and changes in testing practices constrain use of time series methods incorporating post-2020 data. Fifth, RSV and influenza circulation measures are affected by testing practices, changes in participation in laboratory-based surveillance, and diagnostic uncertainty. Sixth, although we used harmonic terms to account for seasonal variation in other factors influencing antibiotic prescription incidence, such as transmission of other respiratory viruses (e.g., human metapneumovirus),^48,49^ residual bias from co-circulating pathogens could persist. Finally, this study is limited to a convenience sample of commercially-insured US children; data may not be broadly representative of US children and findings may not generalize to geographic contexts with differing patterns of seasonal infections^50^ and antibiotic use.^17^

In summary, we found that in the pre-COVID-19 pandemic era, RSV and influenza accounted for 6% and 3% of antibiotic prescriptions dispensed to children, respectively. Beyond disease burden reductions, immunization against these pathogens may reduce antibiotic use, an important step in mitigating antibiotic resistance.

## Author contributions

**LMK:** Conceptualization, methodology, data curation, investigation, and analysis, writing – original draft, writing – review & editing, visualization. **KJB:** Conceptualization, writing – review & editing. **SYT:** Conceptualization, writing – review & editing. **JAL:** Conceptualization, methodology, resources, writing – review & editing, visualization, supervision, project administration.

## Data availability

Data used in this study is proprietary and available for purchase through the Optum Clinformatics™ DataMart.

## Previous presentation

Preliminary results from this study were presented at the 2024 RSVVW conference in Mumbai, India, February 13-16, 2024.

