## Supplemental materials for "Pediatric antibiotic use associated with respiratory syncytial virus and influenza in the United States, 2008-2018"

**Authors:** Laura M King,[1] Katia J Bruxvoort,[2] Sara Y Tartof,[3] Joseph A Lewnard[1]
[1]University of California, Berkeley, School of Public Health, Berkeley, California, USA; [2]University of Alabama at Birmingham, Birmingham, Alabama, USA; [3]Kaiser Permanente Department of Research & Evaluation Southern California, Pasadena, California, USA

**Table S1.** Recommended antibiotic agents by condition

**Table S2.** Diagnosis categories and associated *International Classification of Diseases 9$^{th}$* and *10$^{th}$ revision, Clinical Modification* codes

**Table S3.** Antibiotic class categorization

**Table S4.** Respiratory syncytial virus-, influenza-, and bronchiolitis-associated visits by age category.

**Table S5.** All-cause antibiotic prescription incidence by condition and age group.

**Table S6.** Sensitivity analysis: model-estimated proportion of outpatient antibiotic prescriptions associated with respiratory syncytial virus (RSV) infection in analyses using bronchiolitis visits in children 0-2 years as a proxy for RSV activity.

**Table S7.** Sensitivity analysis: model-estimated proportion of outpatient antibiotic prescriptions associated with influenza virus infection in analyses using bronchiolitis visits in children 0-2 years as respiratory syncytial virus incidence proxy.

**Table S8.** Incidence of respiratory syncytial virus-, influenza-, and bronchiolitis-associated visits by census division.

**Table S9.** Model-estimated proportion of outpatient antibiotic prescriptions associated with respiratory syncytial virus and influenza virus infection, by census division.

**Table S10:** Sensitivity analysis – model-estimated proportion of outpatient antibiotic prescriptions associated with respiratory syncytial virus and influenza infection based on a linear model.

**Table S11.** Sensitivity analysis – model-estimated proportion of outpatient antibiotic prescriptions associated with RSV and influenza infection, when antibiotic definition is expanded to all antibiotics.

**Table S12.** Sensitivity analysis — incidence rates and model-estimated proportion of outpatient antibiotic prescriptions associated with respiratory syncytial virus and influenza virus infection in analyses applying broader inclusion criteria for prescription plan utilization.

**Table S13.** Incidence and proportion of outpatient antibiotic prescriptions associated with respiratory syncytial virus and influenza infection linked to diagnoses for which antibiotics are not indicated, without exclusion of prescriptions linked to other conditions, among children aged 0-17 years, 2008-2018.

**Figure S1.** Predicted and observed antibiotic prescription counts

**Table S1. Recommended antibiotic agents[a] by condition.**

| Condition | Source | Agents[a] |
|---|---|---|
| Acute otitis media | Lieberthal et al. 2013 | Amoxicillin-clavulanate, amoxicillin, cefdinir, cefuroxime, cefpodoxime, ceftriaxone, clindamycin, 3rd generation cephalosporin (cefixime, ceftaxidime, cefotaxime, ceftizoxime, cefpodoxime, ceftriaxone, cefoperazone, ceftibuten, cefdinir, cefditoren) |
| Sinusitis | Wald et al. 2013 | Amoxicillin-clavulanate, amoxicillin, clindamycin, cefixime, linezolid, levofloxacin |
| Pharyngitis | Shulman et al. 2012 | Penicillin V, amoxicillin, penicillin G, cephalexin, cefadroxil, clindamycin, azithromycin, clarithromycin |
| Pneumonia[b] | Bradley et al. 2011 | Amoxicillin, amoxicillin-clavulanate, ampicillin, azithromycin, cefazolin, cefdinir, cefixime, cefotaxime, cefpodoxime, cefprozil, ceftibuten, ceftriaxone, cefuroxime, cephalexin, =floxacin, clarithromycin, clindamycin, doxycycline, erythromycin, levofloxacin, linezolid, moxifloxacin, penicillin, semisynthetic penicillin, vancomycin |

[a] Includes both first-line and alternative agents. Some agents listed recommended in combination. We included all recommended agents in our analysis.

[b] Recommended agents varies by pathogen.

**Table S2. Diagnosis categories and associated *International Classification of Diseases 9th* and *10th revision, Clinical Modification* codes.**

| Diagnosis category | ICD-9-CM codes | ICD-10-CM codes |
|---|---|---|
| RSV[a,b] | 079.6, 466.11, 480.1 | B97.4, J12.1, J20.5, J21.0 |
| Influenza[b,c] | 487*, 488* | J09*-J11*, |
| Acute otitis media[b] | 382*, 384.0*, 384.1 | H66*, H67*, H73-H73.23 |
| Acute exacerbation of COPD[b] | 491.21, 491.22, 493.21, 493.22, 494.1 | J44, J44.0-J44.1 |
| Allergy[b] | 477*, 995.3 | J30*, T78.4, T78.40*, T78.49* |
| Asthma[b] | 493.0*, 493.1*, 493.20, 493.8*, 493.9*, | J45* |
| Bronchiolitis[a,b] | 466.1, 466.19 | J21, J21.1-J21.9 |
| Bronchitis[a,b] | 466.0, 490 | J20-J20.4, J20.6-J20.9, J40 |
| Pharyngitis[b] | 034.0, 462, 463 | J02*, J03* |
| Sinusitis[b] | 461*, 473* | J01*, J32* |
| Pneumonia[a,b,d] | 055.1, 073.0, 480.0, 480.2-486, 507.0, 516.36, 517.1, 770.0, 997.31, 997.32 | A48.1, B01.2, B05.2, B06.81, J12, J12.0, J12.2-J18.9, J69, J69.0, J84.116, J95.851, O29.01*, O74.0, O89.01, P23* |
| Non-suppurative otitis media[b] | 381* | H65*, H68*, H69* |
| Viral URI[b] | 460, 464.0-464.21, 464.4-464.59, 786.2 | J00, J04,*J05.0, J06*, R05 |
| Urinary tract infection[e] | 590.00- 590.11, 590.8-590.9, 595.0-595.2, 595.4, 595.81, 595.89, 595.9, 596.8, 596.81, 599.0, 646.5*, 771.82, | N10, N11- N11.1, N13.6, N30-N30.21, N30.8-N30.91, N39.0, N99.511, N99.521, N99.531, O03.38, O03.88, O04.88, O07.38, O08.83, O23-O23.13, O23.3- O23.43, O86.2*, P39.3 |
| Other bacterial infections[e] | 003.1, 003.21-003.24, 006.3-006.8, 010-033.9, 034.1, 036-041.9, 073.7-073.9, 076-077.0, 077.98, 078.2, 078.3, 078.82, 078.88, 079.88, 079.9, 079.98, 080-084.9, 087-102.9, 104*, 130-131.9, 136.3-137.4, 139.1, 139.8, 254.1, 320-321.1, 321.8-322.9, 323.1-324.9, 326, 349.82, 357.0, 360.00, 360.02, 360.04, 376.02-376.04, 376.12, 379.45, 379.60-379.63, 380.14, 383.0-383.22, 383.81, 383.9, 390-393, 394.1, 395*, 397.1-398.99, 415.12, 420-421.9, 422.92, 424.9*, 440.24, 449, 464.30, 464.31, 475, 478.22, 478.24, 511.1, 513.0, 513.1, 517.3, 519.01, 519.2, 530.86, 536.41, 539.01, 539.81, 540-542, 550.0*, 551*, 564.7, 566-567.81, 567.89, 567.9, 569.5, 569.61, 569.83, 572.0, 572.1, 573.2, 575.4, 576.1, 576.3, 590.2, 597.0, 597.80, 597.89, 598.00, 598.01, 601.0-601.4, 601.9, 603.1, 604*, 607.2, 614.0-614.2, 614.5-616.11, 616.3-616.51, 616.89, 616.9, 634.00-634.02, 635.00-635.02, 636.00-636.02, 637.00-637.02, 638.0, 639.0, 646.60-646.64, 647.00-647.44, 647.80-647.94, 658.4*, 659.3*, 670*, 673.3*, 675.1*, 695.81, 711.00-711.19, 711.9*, 728.0, 728.86, 730.00-730.29, 730.80-730.99, 760.2, 770.12, 771.2-771.4, 771.81, 771.83, 771.89, 777.5*, 777.6, 785.4, 785.52, 788.7, 790.7, 795.3*, 958.3, 995.91, 995.92, 996.6*, 997.62, 998.02, 998.51, 998.59, 999, 999.31, 999.32, 999.34, 999.39, V01.0, V01.1, V01.6, V01.81, V01.83-V01.89, V01.9, V02.8, V09*, V73.88, V73.98 | A01, A01.0, A01.01-A01.05, A02, A02.1, A02.2, A02.21-A02.29, A06, A06.4-A06.82, A15*-A44*, A48, A48.0, A48.2-A59.9, A63, A63.8, A64*-A66*, A68*-A79*, B47, B47.1-B47.9, B50*-B54*, B58-B60.0, B60.2, B90*, B92-B94.0, B94.8-B96.89, B99*, D57.01, D57.211, D57.411, D57.811, D73.3, E08.52, E09.52, E10.52, E11.52, E13.52, E32.1, G00-G03.1, G03.8-G04.02, G04.2-G07, G92, H05, H05.0, H05.02-H05.049, H05.1, H05.12*, H44, H44.0*, H57.01, H59.4-H59.43, H60.2*, H70*, H75*, I00-I09.9, I30*, I32-I33.9, I38, I39, I40.0, I70.26*, I70.36*, I70.46*, I70.56*, I70.66*, I70.76*, I73.01, I76, I96, J05, J05.1*, J36, J39.0, J39.1, J47-J47.1, J65, J80, J85-J86.9, J98.5, J98.51, K35-K37, K40.1*, K40.4*, K41.1*, K41.4*, K42.1, K43.1, K43.4, K43.7, K44.1, K45.1, K46.1, K50.014, K50.114, K50.814, K50.914, K51.014, K51.214, K51.314, K51.414, K51.514, K51.814, K51.914, K55.3*, K57.0*, K57.2*, K57.4*, K57.8*, K59.3*, K61*, K63.0, K63.1, K65-K65.3, K65.8, K65.9, K68-K68.19, K75.0, K80.3*, K82.2, K82.A*, K83.0*, K83.2, K85.02, K85.12, K85.22, K85.32, K85.82, K85.92, K90.81, K94.02, K94.12, K94.22, K94.32, K95.01, K95.81, L00, M00-M01.X9, M02.3*, M46.2-M46.39, M46.5*, M60.0*, M65.0-M65.19, M71.0 -M71.19, M72.6, M86*, N15.1, N34-N34.2, N41-N41.3, N43.1, N45,*, N48.2, N48.21, N49.3, N70-N73, N73.3-N74, N75.1, N76-N76.6, N77*, N98.0, O03.0, O03.37, O03.5, O03.87, O04.5, O04.87, O07.0, O07.37, O08.0, O08.82, O23.2*, O23.5*. O23.9*, O41.1*, O75.3, O85-O86.19, O86.8*, O88.3*, O91.1*, O98-O98.33, O98.8-O98.93, O99.8, O99.82-O99.835, P00.2, P02.7*, P24.0, P24.01, P36-P37.2, P37.8-P39, P39.2, P39.8, P39.9, P58.2, P77*, P78.0, R36, R36.0, R36.9, R65, R65.2-R65.21, R78.81, T80.2-T80.211S, T80.218-T80.29XS, T81.12*, T81.4*, T82.6-T82.7XXS, T83.5-T83.6XXS, T84.5-T84.7XXS, T85, T85.7*, T86.03, T86.13, T86.23, T86.33, T86.43, T86.812, T86.832, T86.852, T86.892, T86.93, T87.4*, Z11.3, Z20, Z20.1, Z20.2, Z20.8, Z20.81*, Z20.89, Z20.9, Z71.84 |

| Diagnosis category | ICD-9-CM codes | ICD-10-CM codes |
|---|---|---|
| Gastrointestinal infections | 001.0-003.0, 003.20, 003.29-006.2, 006.9-009.3, 522.0, 522.5, 522.7, 526.5, 527.2-, 527.3, 533*, 562.01, 562.03, 562.11, 562.13, 569.7*, 569.87, 577.0, 579.1, 579.2, 779.32, 787.04, 787.6*, 787.91, 789.1, 789.2 | A00, A00.0-A00.1, A00.9, A01.00, A01.09, A01.1-A01.4, A02.0, A02.20, A02.8-A05.9, A06.0-A06.3, A06.89-A09, K04-K04.1, K04.6-K04.7, K11, K11.2-K11.3, K11.8, K11.9, K27*, K51.5-K51.513, K51.518-K51.519, K57, K57.1, K57.12, K57.13, K57.3, K57.32, K57.33, K57.5, K57.52, K57.53, K57.9, K57.92, K57.93, K67, K85-K85.01, K85.1-K85.11, K85.2-K85.21, K85.3-K85.31, K85.8-K85.81, K85.9-K85.91, K90.1-K90.2, K91.85-K91.858, M27.3, M27.51, P78.1, P78.82, R11.12-R11.14, R15-R16.2, R19.7, Z20.0-Z20.09 |
| Skin and soft tissue infections | 035, 103*, 110-111.9, 360.01, 360.03, 360.11-360.13, 360.19, 363.00-363.21, 364.00-364.03, 364.05-364.21, 364,3, 370.00-370.22, 370.31, 370.40-370.52, 370.55, 370.59, 370.8, 370.9, 372.00-372.04, 372.10-372.12, 372.15-372.21, 372.30-372.33, 372.39, 373.00-373.2, 373.6, 375.01, 375.02, 375.30-375.42, 376.01, 376.13, 380.00-380.13, 380.15, 380.16, 380.22, 380.23, 457.2, 478.21, 478.71, 528.0-528.3, 611.0, 611.5, 614.3, 614.4, 675.0*, 675.20-675.94, 680.0-686.09, 686.8, 686.9, 694.1, 694.3, 695.3, 705.82, 705.83, 706.0, 706.1, 723.6, 729.30, 729.39, 771.5, 771.6, 910.1, 910.3, 910.5, 910.7, 910.9, 911.1, 911.3, 911.5, 911.7, 911.9, 912.1, 912.3, 912.5, 912.7, 912.9, 913.1, 913.3, 913.5, 913.7, 913.9, 914.1, 914.3, 914.5, 914.7, 914.9, 915.1, 915.3, 915.5, 915.7, 915.9, 916.1, 916.3, 916.5, 916.7, 916.9, 917.1, 917.3, 917.5, 917.7, 917.9, 919.1, 919.3, 919.5, 919.7, 919.9, 999.33, E90.60-E90.63, E90.65, E92.83, E96.87 | A46, A67*, B35*-B36*, H00-H01.02B, H04-H04.029, H04.3-H04.429, H05.01*, H10-H10.029, H10.2, H10.22-H10.409, H10.42-H10.439, H10.5-H10.509, H10.52*, H10.8, H10.82-H10.9, H16-H16.119, H16.14-H16.209, H16.25*, H16.29-H16.329, H16.39*, H16.8, H16.9, H20-H20.039, H20.05-H20.13, H20.8-H20.819, H20.9, H21.33*, H33.12*, H44.1*, H60-H60.13, H60.3*, H60.6-H61.039, H62-H62.43, H94-H94.03, I89.1, J34.0, J95.02, K12-K12.39, L01-L08.9, L66.3, L70*, L71, L71.8, L71.9, L73*, L88, M79.3, N48.22, N61*, N73.0-N73.2, O91-O91.03, O91.2-O91.23, P39.0, P39.1, P39.4, S00.07, S00.07XA, S00.27, S00.271, S00.271A, S00.272, S00.72A, S00.279, S00.279A, S00.37, S00.37XA, S00.47, S00.471, S00.471A, S00.472, S00.472A, S00.479, S00.479A, S00.57, S00.571, S00.571A, S00.572, S00.572A, S00.87, S00.87XA, S00.97, S00.97XA, S01.05, S01.05XA, S01.15, S01.151, S01.151A, S01.152, S01.152A, S01.159, S01.159A, S01.25, S01.25XA, S01.35, S01.351, S01.351A, S01.352, S01.352A, S01.359, S01.359A, S01.45, S01.451, S01.451A, S01.452, S01.452A, S01.459, S01.459A, S01.55, S01.551, S01.551A, S01.552, S01.552A, S01.85, S01.85XA, S01.95, S01.95XA, S10.17, S10.17XA, S10.87, S10.87XA, S10.97, S10.97XA, S11.015, S11.015A, S11.025, S11.025A, S11.035, S11.035A, S11.15, S11.15XA, S11.25, S11.25XA, S11.85, S11.95, S11.95XA, S20.17, S20.171, S20.171A, S20.172, S20.172A, S20.179, S20.179A, S20.37, S20.371, S20.371A, S20.372, S20.372A, S20.379, S20.379A, S20.47, S20.471, S20.471A, S20.472, S20.472A, S20.479, S20.479A, S20.97, S20.97XA, S21.05, S21.051, S21.051A, S21.052, S21.052A, S21.059, S21.059A, S21.15, S21.151, S21.151A, S21.152, S21.152A, S21.159, S21.159A, S21.25, S21.251, S21.251A, S21.252, S21.252A, S21.259, S21.259A, S21.35, S21.351, S21.351A, S21.352, S21.352A, S21.359, S21.359A, S21.45, S21.451, S21.451A, S21.452, S21.452A, S21.459, S21.459A, S21.95, S21.95XA, S30.87, S30.870, S30.870A, S30.871, S30.871A, S30.872, S30.872A, S30.873, S30.873A, S30.874, S30.874A, S30.875, S30.875A, S30.876, S30.876A, S30.877, S30.877A, S31.05, S31.050, S31.050A, S31.051, S31.051A, S31.15, S31.150, S31.150A, S31.151, S31.151A, S31.152, S31.152A, S31.153, S31.153A, S31.154, S31.154A, S31.155, S31.155A, S31.159, S31.159A, S31.25, S31.25XA, S31.35, S31.35XA, S31.45, S31.45XA, S31.55, S31.551, S31.551A, S31.552, S31.552A, S31.65, S31.650, S31.650A, S31.651, S31.651A, S31.652, S31.652A, S31.653, S31.653A, S31.654, S31.654A, S31.655, |

| Diagnosis category | ICD-9-CM codes | ICD-10-CM codes |
|---|---|---|
| | | S31.655A, S31.659, S31.659A, S31.805, S31.805A, S31.815, S31.815A, S31.825, S31.825A, S31.835, S31.835A, S40.27, S40.271, S40.271A, S40.272, S40.272A, S40.279, S40.279A, S40.87, S40.871, S40.871A, S40.872, S40.872A, S40.879, S40.879A, S41.05, S41.051, S41.051A, S41.052, S41.052A, S41.059, S41.059A, S41.15, S41.151, S41.151A, S41.152, S41.152A, S41.159, S41.159A, S50.37, S50.371, S50.371A, S50.372, S50.372A, S50.379, S50.379A, S50.87, S50.871, S50.871A, S50.872, S50.872A, S50.879, S50.879A, S51.05, S51.051, S51.051A, S51.052, S51.052A, S51.059, S51.059A, S51.85, S51.851, S51.851A, S51.852, S51.852A, S51.859, S51.859A, S60.37, S60.371, S60.371A, S60.372, S60.372A, S60.379, S60.379A, S60.47, S60.470, S60.470A, S60.471, S60.471A, S60.472, S60.472A, S60.473, S60.473A, S60.474, S60.474A, S60.475, S60.475A, S60.476, S60.476A, S60.477, S60.477A, S60.478, S60.478A, S60.479, S60.479A, S60.57, S60.571, S60.571A, S60.572, S60.572A, S60.579, S60.579A, S60.87, S60.871, S60.871A, S60.872, S60.872A, S60.879, S60.879A, S61.05, S61.051, S61.051A, S61.052, S61.052A, S61.059, S61.059A, S61.15, S61.151, S61.151A, S61.152, S61.152A, S61.159, S61.159A, S61.25, S61.250, S61.250A, S61.251, S61.251A, S61.252, S61.252A, S61.253, S61.253A, S61.254, S61.254A, S61.255, S61.255A, S61.256, S61.256A, S61.257, S61.257A, S61.258, S61.258A, S61.259, S61.259A, S61.35, S61.350, S61.350A, S61.351, S61.351A, S61.352, S61.352A, S61.353, S61.353A, S61.354, S61.354A, S61.355, S61.355A, S61.356, S61.356A, S61.357, S61.357A, S61.358, S61.358A, S61.359, S61.359A, S61.45, S61.451, S61.451A, S61.452, S61.452A, S61.459, S61.459A, S61.55, S61.551, S61.551A, S61.552, S61.552A, S61.559, S61.559A, S70.27, S70.271, S70.271A, S70.272, S70.272A, S70.279, S70.279A, S70.37, S70.371, S70.371A, S70.372, S70.372A, S70.379, S70.379A, S71.05, S71.051, S71.051A, S71.052, S71.052A, S71.059, S71.059A, S71.15, S71.151, S71.151A, S71.152, S71.152A, S71.159, S71.159A, S80.27, S80.271, S80.271A, S80.272, S80.272A, S80.279, S80.279A, S80.87, S80.871, S80.871A, S80.872, S80.872A, S80.879, S80.879A, S81.05, S81.051, S81.051A, S81.052, S81.052A, S81.059, S81.059A, S81.85, S81.851, S81.851A, S81.852, S81.852A, S81.859, S81.859A, S90.47, S90.471, S90.471A, S90.472, S90.472A, S90.473, S90.473A, S90.474, S90.474A, S90.475, S90.475A, S90.476, S90.476A, S90.57, S90.571, S90.571A, S90.572, S90.572A, S90.579, S90.579A, S90.87, S90.871, S90.871A, S90.872, S90.872A, S90.879, S90.879A, S91.05, S91.051, S91.051A, S91.052, S91.052A, S91.059, S91.059A, S91.15, S91.151, S91.151A, S91.152, S91.152A, S91.153, S91.153A, S91.154, S91.154A, S91.155, S91.155A, S91.156, S91.156A, S91.159, S91.159A, S91.25, S91.251, S91.251A, S91.252, S91.252A, S91.253, S91.253A, S91.254, S91.254A, S91.255, S91.255A, S91.256, S91.256A, S91.259, S91.259A, S91.35, S91.351, |

| Diagnosis category | ICD-9-CM codes | ICD-10-CM codes |
|---|---|---|
| | | S91.351A, S91.352, S91.352A, S91.359, S91.359A, T80.212*, T86.822, T86.842, W50, W50.3, W50.3XXA, W53.01, W53.01XA, W53.11, W53.11XA, W53.21, W53.21XA, W53.81, W53.81XA, W54.0, W54.0XXA, W55.01, W55.01XA, W55.11, W55.11XA, W55.21, W55.21XA, W55.31, W55.31XA, W55.41, W55.41XA, W55.51, W55.51XA, W55.81, W55.81XA, W56.01, W56.01XA, W56.11, W56.11XA, W56.21, W56.21XA, W56.31, W56.31XA, W56.41, W56.41XA, W56.51, W56.51XA, W56.81, W56.81XA, W58.01, W58.01XA, W58.11, W58.11XA, W59.01, W59.01XA, W59.11, W59.11XA, W59.21, W59.21XA, W59.81, W59.81XA, W61.01, W61.01XA, W61.11, W61.11XA, W61.2, W61.21, W61.21XA, W61.51, W615.1XA, W61.61, W61.61XA, W61.91, W61.91XA, Y04.1, Y04.1XXA |

Abbreviations: ICD-9-CM -- *International Classification of Diseases 9th revision, Clinical Modification;* ICD-10-CM -- *International Classification of Diseases 10th revision, Clinical Modification;* RSV – respiratory syncytial virus; COPD – chronic obstructive pulmonary disorder.

* Includes all child codes beginning with the numeric sequence.

[a] Categorized as lower respiratory tract infection (LRTI).

[b] Categorized as acute respiratory infection (ARI).

[c] Includes acute otitis media and pneumonia due to influenza.

[d] Includes viral pneumonia except pneumonia related to influenza, which is captured under influenza.

[e] Used to exclude antibiotic prescriptions from being attributed to ARI-related diagnoses when ARI and UTI or ARI and other bacterial infections are both present for a single visit.

**Table S3. Antibiotic class categorization.**

| Class | Antibiotic agent[a] |
|---|---|
| Penicillin | Amoxicillin, ampicillin, penicillin G, penicillin V, carbenicillin, dicloxacillin |
| Extended-spectrum beta-lactam | Amoxicillin-clavulanate, am*picillin-sulbactam* |
| Macrolide | Azithromycin, clarithromycin, erythromycin, *erythromycin-sulfisoxazole* |
| Cephalosporin | Cefdinir, cefuroxime, cefpodoxime, ceftriaxone, cefixime, cefotaxime, cefditoren, ceftibuten, cephalexin, cefadroxil, cefazolin, cefprozil, *cefaclor, cefepime, ceftazidime* |
| Tetracycline | Doxycycline, *minocycline, tetracycline, demecylocycline* |
| Fluoroquinolone | Levofloxacin, moxifloxacin, ciprofloxacin, *delafloxacin, gatifloxacin, gemifloxacin, norfloxacin, oflaxacin, besifloxacin* |
| Other | Clindamycin, linezolid, vancomycin, *amikacin, chloramphenicol, colistimethate, colistin, fosfomycin, furazolidone, gentamycin, lincomycin, loracarbef, meropenem, methenamine, nitrofurantoin, rifaximin, secnidazole, sulfadiazine, sulfamethoxazole, sulfamethoxazole-trimethoprim, sulfanilamid, sulfisoxazole, tedizolid, telithromycin, trimethoprim,* |

[a] Includes agents with >1 dispensing event in the Clinformatics database. Both agents classified as respiratory and non-respiratory included. Agents not categorized as respiratory (**Table S1**) are italicized. Only agents categorized as respiratory included in primary analyses, all agents included in sensitivity analysis.

**Table S4. Respiratory syncytial virus-, influenza-, and bronchiolitis-linked visits by age category.**

| Age category | Visit[a] incidence per 1000 person-years (95% CI)[b] | | |
|---|---|---|---|
| | *RSV* | *Influenza* | *Bronchiolitis* |
| 0-2 years | 207 (180, 236) | 111 (91, 132) | 527 (483, 573) |
| 3-5 years | 24 (15, 34) | 139 (116, 162) | 63 (48, 79) |
| 6-9 years | 6 (2, 11) | 151 (128, 176) | 15 (8, 24) |
| 10-13 years | 3 (0, 8) | 124 (103, 146) | 8 (3, 14) |
| 14-17 years | 3 (0, 6) | 83 (66, 102) | 5 (1, 10) |
| Total | 53 (39, 68) | 121 (100, 143) | 101 (82, 121) |

Abbreviations: CI – confidence interval; RSV – respiratory syncytial virus.

[a] All visits with any diagnostic code for RSV, influenza, or bronchiolitis as detailed in **Table S2.** We additionally included visits with ICD-9-CM code 466.11 or ICD-10-CM code J21.0 (bronchiolitis due to RSV) in the definition of bronchiolitis visits.

[b] Obtained by fitting number of antibiotic prescriptions divided by person-years to a Poisson distribution, sampling 100,000 random draws, and obtaining the median, 2.5$^{th}$ and 97.5$^{th}$ percentile values.

**Table S5. All-cause antibiotic prescription incidence by condition and age group.**

| Linked diagnosis[b] | | Antibiotic prescription incidence per 1000 person-years (95% CI)[a] | | | | | |
|---|---|---|---|---|---|---|---|
| | | *All children* | *0-2 years* | *3-5 years* | *6-9 years* | *10-13 years* | *14-17 years* |
| Any[c] | | 1,161 (1,096, 1,228) | 1,834 (1,752, 1,917) | 1,469 (1,396, 1,544) | 1,129 (1,065, 1,195) | 846 (790, 902) | 897 (841, 956) |
| Respiratory syndromes | | | | | | | |
| | Any ARI[d,e] | 775 (723, 830) | 1,394 (1,323, 1,467) | 1,081 (1,018, 1,145) | 800 (747, 856) | 554 (509, 600) | 412 (373, 452) |
| | Any LRTI[e,f] | 99 (81, 119) | 149 (126, 173) | 144 (121, 168) | 101 (83, 122) | 76 (60, 93) | 62 (48, 78) |
| Specific condition | | | | | | | |
| | Pneumonia[e,g] | 29 (20, 41) | 41 (29, 54) | 50 (37, 65) | 34 (24, 47) | 21 (13, 31) | 12 (6, 19) |
| | AOM[e] | 275 (244, 308) | 864 (808, 922) | 446 (406, 488) | 210 (183, 239) | 103 (84, 123) | 55 (41, 70) |
| | Sinusitis[e] | 147 (124, 171) | 153 (130, 178) | 170 (145, 196) | 136 (114, 159) | 140 (118, 164) | 145 (123, 170) |
| | Pharyngitis[e] | 229 (201, 260) | 140 (117, 163) | 275 (243, 308) | 336 (301, 372) | 233 (204, 263) | 151 (128, 176) |
| | Bronchiolitis[h,i] | 4 (1, 8) | 17 (9, 25) | 5 (1, 10) | 2 (0, 5) | 1 (0, 3) | 0 (0, 2) |
| | Bronchitis[i,j] | 42 (30, 55) | 41 (29, 54) | 60 (45, 75) | 45 (33, 59) | 37 (26, 49) | 32 (22, 44) |
| | Nonsupp. OM[i] | 32 (21, 43) | 97 (79, 117) | 52 (38, 66) | 24 (15, 35) | 12 (6, 19) | 7 (2, 12) |
| | Allergy[i] | 11 (5, 18) | 8 (3, 14) | 13 (7, 21) | 13 (7, 21) | 11 (5, 18) | 9 (4, 15) |
| | Asthma[i] | 16 (9, 25) | 14 (7, 22) | 22 (13, 31) | 20 (12, 29) | 16 (9, 24) | 11 (5, 18) |
| | Viral URI (excl. RSV and influenza)[i] | 75 (59, 93) | 129 (108, 152) | 109 (90, 130) | 72 (56, 89) | 54 (40, 69) | 45 (33, 59) |
| | Any RSV diagnosis[i] | 1 (0, 4) | 6 (2, 11) | 1 (0, 4) | 0 (0, 2) | 0 (0, 1) | 0 (0, 1) |
| | Any influenza diagnosis[i,k] | 4 (1, 9) | 3 (0, 7) | 5 (1, 10) | 5 (2, 11) | 4 (1, 9) | 3 (0, 7) |

Abbreviations: CI – confidence interval; ARI – acute respiratory infection; LRTI – lower respiratory tract infection; AOM – acute otitis media; Non-supp. OM – nonsuppurative otitis media; URI – upper respiratory infection; RSV – respiratory syncytial virus.

[a] Obtained by fitting number of antibiotic prescriptions divided by person-years to a Poisson distribution and sampling 100,000 random draws.

[b] Based on ICD-9-CM and ICD-10-CM codes (**Table S3**) assigned at the most recent outpatient visit within a 4-day window. More than one diagnosis can be associated with an antibiotic prescription.

[c] Includes antibiotic prescriptions for any diagnosis.

[d] Includes diagnoses of pneumonia, AOM, sinusitis, pharyngitis, bronchiolitis, bronchitis, non-suppurative otitis media, viral upper respiratory infection, RSV, influenza, and acute exacerbation of chronic bronchitis/obstructive pulmonary disorder (accounts for <0.04% of antibiotic prescriptions).

[e] Antibiotic prescriptions with concurrent diagnoses of urinary tract infection and other bacterial infections excluded.

[f] Includes diagnoses of bronchitis, bronchiolitis, and pneumonia excluding diagnosis codes for these conditions due to RSV or influenza.

[g] Excludes pneumonia due to influenza and RSV but includes other viral pneumonia. Among pneumonia prescriptions, 2.4% had a viral pneumonia diagnosis with no other pneumonia diagnosis.

[h] Excludes diagnosis codes for bronchiolitis due to RSV.

[i] Antibiotic prescriptions with concurrent diagnoses of urinary tract infection, other bacterial infections, pneumonia, AOM, sinusitis, pharyngitis, gastrointestinal infections, and skin and soft tissue infections excluded.

[j] Excludes diagnosis codes for bronchitis due to RSV.

[k] Includes all diagnoses of influenza, including influenza and pneumonia and influenza and AOM.

**Table S6. Sensitivity analysis: model-estimated proportion of outpatient antibiotic prescriptions associated with respiratory syncytial virus (RSV) infection in analyses using bronchiolitis visits in children 0-2 years as a proxy for RSV activity.[a]**

| Linked diagnosis[b] | | Percent of antibiotic prescriptions associated with RSV infection (95% CI) | | | | | |
|---|---|---|---|---|---|---|---|
| | | *All children* | *0-2 years* | *3-5 years* | *6-9 years* | *10-13 years* | *14-17 years* |
| Any[c] | | 8.0 (6.8, 9.3) | 11.2 (8.0, 14.4) | 10.0 (6.7, 13.4) | 6.9 (3.6, 10.2) | 6.4 (4.0, 8.7) | 4.3 (2.8, 5.8) |
| Respiratory syndromes | | | | | | | |
| | Any ARI[d,e] | 9.2 (7.5, 10.9) | 11.5 (7.9, 15.1) | 11.4 (7.4, 15.4) | 7.7 (3.4, 11.9) | 7.7 (3.9, 11.5) | 7.3 (4.3, 10.4) |
| | Any LRTI[e,f] | 17.0 (15.2, 18.8) | 32.2 (29.1, 35.2) | 21.0 (17.0, 24.9) | 12.0 (7.3, 16.4) | 8.3 (3.6, 12.9) | 7.7 (3.4, 11.8) |
| Specific conditions | | | | | | | |
| | Pneumonia[e,g] | 15.4 (13.6, 17.2) | 25.7 (22.5, 28.7) | 23.0 (19.3, 26.5) | 11.9 (7.6, 15.9) | 6.6 (1.9, 11.1) | 7.5 (2.8, 12.0) |
| | AOM[e] | 12.3 (10.7, 14.0) | 12.3 (8.6, 16.0) | 16.0 (12.0, 19.8) | 14.0 (9.9, 17.8) | 10.3 (6.6, 13.9) | 8.0 (4.7, 11.3) |
| | Sinusitis[e] | 6.3 (4.7, 8.0) | 3.9 (0.1, 7.6) | 6.2 (2.2, 10.2) | 5.8 (1.6, 9.9) | 7.9 (4.1, 11.6) | 8.5 (5.3, 11.6) |
| | Pharyngitis[e] | 4.8 (2.9, 6.7) | 4.3 (0.5, 7.9) | 3.6 (-0.8, 7.9) | 3.2 (-1.8, 7.9) | 6.2 (1.6, 10.7) | 7.0 (3.4, 10.5) |
| | Bronchiolitis[h,i] | 26.7 (24.2, 29.1) | 40.3 (37.1, 43.4) | 29.8 (24.8, 34.6) | 14.0 (5.8, 21.5) | 8.6 (-2.3, 18.3) | 4.2 (-8.4, 15.3) |
| | Bronchitis[i,j] | 11.3 (9.0, 13.6) | 15.5 (11.1, 19.7) | 15.7 (10.9, 20.4) | 10.3 (4.7, 15.6) | 8.5 (2.7, 14.0) | 6.4 (0.9, 11.6) |
| | Non-suppurative otitis media[i] | 10.5 (8.8, 12.2) | 11.1 (7.5, 14.6) | 13.3 (9.4, 17.0) | 11.4 (7.5, 15.2) | 8.6 (4.9, 12.3) | 6.6 (2.6, 10.4) |
| | Allergy[i] | 5.5 (3.8, 7.3) | 2.3 (-2.5, 7.0) | 7.6 (3.8, 11.4) | 4.2 (0.2, 8.0) | 6.7 (3.0, 10.3) | 4.2 (0.7, 7.6) |
| | Asthma[i] | 9.6 (7.8, 11.5) | 15.5 (11.7, 19.2) | 11.0 (6.8, 15.0) | 9.0 (4.6, 13.2) | 7.7 (3.3, 12.0) | 5.1 (0.9, 9.2) |
| | Viral URI[i] (excl. RSV and influenza) | 11.0 (9.1, 12.8) | 13.1 (9.4, 16.8) | 13.5 (9.3, 17.6) | 9.4 (4.6, 13.9) | 9.9 (5.3, 14.4) | 8.8 (4.6, 12.9) |
| | Any RSV diagnosis[i] | 51.9 (49.6, 54.2) | 63.3 (60.7, 65.8) | 55.3 (50.7, 59.5) | 26.0 (14.8, 35.3) | 2.4 (-20.3, 20.2) | -0.5 (-26.2, 18.9) |
| | Any influenza diagnosis[i,k] | 25.3 (21.9, 28.6) | 23.1 (14.3, 31.0) | 29.0 (21.2, 36.1) | 26.2 (18.0, 33.4) | 20.0 (12.8, 26.8) | 26.1 (18.4, 33.2) |

Abbreviations: CI – confidence interval; RSV – respiratory syncytial virus; ARI – acute respiratory infection; LRTI – lower respiratory tract infection; AOM – acute otitis media; Non-supp. OM – nonsuppurative otitis media; URI – upper respiratory infection.

[a] Primary analysis used the percent of samples positive for RSV measured from laboratory data from NREVSS lagged one week behind antibiotic prescriptions as the RSV transmission proxy.

[b] Based on ICD-9-CM and ICD-10-CM codes (**Table S3**) assigned at the most recent outpatient visit within a 4-day window. More than one diagnosis can be associated with an antibiotic prescription.

[c] Includes antibiotic prescriptions for any diagnosis.

[d] Includes diagnoses of pneumonia, sinusitis, pharyngitis, AOM, asthma/allergy, nonsuppurative otitis media, bronchitis/bronchiolitis, influenza, RSV, acute exacerbation of chronic bronchitis/obstructive pulmonary disorder (accounts for <0.04% of antibiotic prescriptions), and viral upper respiratory infection.

[e] Antibiotic prescriptions with concurrent diagnoses of urinary tract infection and other bacterial infections excluded.

[f] Includes diagnoses of bronchitis, bronchiolitis, and pneumonia excluding diagnosis codes for these conditions due to RSV or influenza.

[g] Excludes pneumonia due to influenza and RSV but includes other viral pneumonia. Among pneumonia prescriptions, 2.4% had a viral pneumonia diagnosis with no other pneumonia diagnosis.

[h] Excludes diagnosis codes for bronchiolitis due to RSV.

[i] Antibiotic prescriptions with concurrent diagnoses of urinary tract infection, other bacterial infections, pneumonia, AOM, sinusitis, pharyngitis, gastrointestinal infections, and skin and soft tissue infections excluded.

[j] Excludes diagnosis codes for bronchitis due to RSV.

[k] Includes all diagnoses of influenza, including influenza and pneumonia and influenza and AOM.

**Table S7. Sensitivity analysis: model-estimated proportion of outpatient antibiotic prescriptions associated with influenza virus infection in analyses using bronchiolitis visits in children 0-2 years as respiratory syncytial virus incidence proxy.[a,b]**

| Age group | Percent of antibiotic prescriptions associated with infection (95% CI) |
|---|---|
| 0-2 years | 0.3 (-0.9, 1.4) |
| 3-5 years | 1.3 (0.2, 2.4) |
| 6-9 years | 3.4 (2.4, 4.4) |
| 10-13 years | 4.1 (3.4, 4.8) |
| 14-17 years | 3.3 (2.8, 3.7) |
| All children | 3.1 (2.8, 3.5) |

Abbreviations: CI – confidence interval

[a] Primary analysis used the percent of samples positive for RSV measured from laboratory data from NREVSS lagged one week behind antibiotic prescriptions as the RSV transmission proxy.

[b] Includes antibiotic prescriptions for any diagnosis.

**Table S8. Incidence of respiratory syncytial virus-, influenza-, and bronchiolitis-associated visits by census division.**

| Census division | Visit incidence[a] per 1000 person-years (95% CI)[b] | | |
|---|---|---|---|
| | *RSV* | *Influenza* | *Bronchiolitis* |
| East North Central | 43 (31, 57) | 80 (63, 98) | 83 (66, 101) |
| East South Central | 108 (89, 129) | 219 (194, 248) | 135 (113, 158) |
| Middle Atlantic | 51 (38, 66) | 97 (78, 117) | 113 (93, 135) |
| Mountain | 69 (54, 86) | 89 (71, 108) | 116 (96, 138) |
| New England | 79 (62, 97) | 97 (78, 117) | 128 (107, 151) |
| Pacific | 35 (24, 48) | 63 (48, 79) | 74 (58, 92) |
| South Atlantic | 43 (31, 56) | 124 (103, 147) | 95 (76, 114) |
| West North Central | 72 (56, 89) | 105 (85, 125) | 109 (90, 131) |
| West South Central | 60 (46, 76) | 191 (165, 219) | 114 (94, 136) |
| Total | 53 (39, 68) | 121 (100, 143) | 101 (82, 121) |

Abbreviations: CI – confidence interval; RSV – respiratory syncytial virus.

[a] All visits with any diagnostic code for RSV, influenza, or bronchiolitis as detailed in **Table S3.** We additionally included visits with ICD-9-CM code 466.11 or ICD-10-CM code J21.0 (bronchiolitis due to RSV) in the definition of bronchiolitis visits.

[b] Obtained by fitting number of antibiotic prescriptions divided by person-years to a Poisson distribution, sampling 100,000 random draws, and obtaining the median, 2.5$^{th}$ and 97.5$^{th}$ percentile values.

**Table S9. Model-estimated proportion of outpatient antibiotic prescriptions associated with respiratory syncytial virus and influenza virus infection, by census division.**

| Census division | Percent of antibiotic prescriptions associated with infection (95% CI) | | Antibiotic prescription incidence per 1000 person-years (95% CI) | |
|---|---|---|---|---|
| | *RSV* | *Influenza* | *RSV* | *Influenza* |
| East North Central | 3.4 (-1.0, 7.4) | 3.2 (2.2, 4.3) | 38.8 (-11.2, 84.3) | 36.6 (24.8, 48.9) |
| East South Central | 5.0 (1.1, 8.6) | 4.1 (2.9, 5.3) | 71.7 (16.0, 122.9) | 59.2 (42.0, 76.9) |
| Middle Atlantic | 6.7 (2.5, 10.6) | 3.6 (2.5, 4.7) | 76.2 (28.3, 120.7) | 40.8 (27.9, 54.2) |
| Mountain | 3.6 (-0.7, 7.5) | 4.0 (2.8, 5.1) | 36.6 (-6.7, 76.5) | 40.4 (28.7, 52.6) |
| New England | 5.2 (1.4, 8.6) | 3.0 (2.1, 4.0) | 52.8 (14.2, 88.4) | 30.8 (21.2, 40.8) |
| Pacific | 4.4 (0.2, 8.3) | 3.5 (2.3, 4.7) | 41.4 (2.2, 77.1) | 32.9 (21.6, 44.6) |
| South Atlantic | 7.3 (0.6, 13.4) | 4.2 (2.9, 5.5) | 85.1 (6.8, 157.0) | 48.4 (33.2, 63.9) |
| West North Central | 3.7 (-2.2, 9.0) | 2.7 (1.7, 3.7) | 43.9 (-26.7, 108.3) | 31.7 (19.9, 44.0) |
| West South Central | 5.3 (0.6, 9.6) | 4.6 (3.3, 5.8) | 71.5 (8.0, 129.8) | 61.7 (44.6, 79.2) |

Abbreviations: CI – confidence interval; RSV – respiratory syncytial virus.

**Table S10. Sensitivity analysis – model-estimated proportion of outpatient antibiotic prescriptions associated with respiratory syncytial virus and influenza infection based on a linear model.**

| **Age group** | **Associated fraction based on linear model,[a] % (95% CI)** | |
|---|---|---|
| | *RSV* | *Influenza* |
| 0-2 years | 8.9 (7.1, 10.9) | 1.3 (0.6, 2.0) |
| 3-5 years | 8.7 (6.9, 10.5) | 1.9 (1.3, 2.5) |
| 6-9 years | 5.2 (3.3, 7.1) | 3.5 (2.9, 4.1) |
| 10-13 years | 3.8 (2.2, 5.5) | 4.2 (3.7, 4.6) |
| 14-17 years | 2.5 (1.5, 3.6) | 3.5 (3.2, 3.8) |
| All children [b] | 6.8 (6.0, 7.7) | 3.1 (2.8, 3.3) |

Abbreviations: CI – confidence interval; RSV – respiratory syncytial virus.

[a] Model of the incidence of antibiotic prescriptions (no. prescriptions/person-weeks) as a linear function of census division, 13-valent pneumococcal conjugate vaccine period, 1-week lagged percent of positive RSV tests, influenza incidence by age category, 12-, 6-, 4- and 3-month harmonics, and week secular trend term.

[b] The model for all children also controlled for age category.

**Table S11: Sensitivity analysis – model-estimated proportion of outpatient antibiotic prescriptions associated with RSV and influenza infection, when antibiotic definition is expanded to all antibiotics.**

| Age group | Fraction associated with each infection for all agents[a] % (95% CI) | |
|---|---|---|
| | *RSV* | *Influenza* |
| 0-2 years | 8.4 (5.7, 11.1) | 0.9 (-0.2, 2.0) |
| 3-5 years | 8.3 (5.5, 11.1) | 1.7 (0.7, 2.8) |
| 6-9 years | 5.6 (3.0, 8.2) | 3.6 (2.7, 4.5) |
| 10-13 years | 4.0 (2.3, 5.7) | 4.1 (3.5, 4.6) |
| 14-17 years | 2.2 (1.0, 3.3) | 3.0 (2.6, 3.4) |
| All children | 6.1 (5.1, 7.1) | 3.3 (3.0, 3.7) |

Abbreviations: CI – confidence interval; RSV – respiratory syncytial virus.
[a] Includes all agents listed in **Table S2**.

**Table S12. Sensitivity analysis — incidence rates and model-estimated proportion of outpatient antibiotic prescriptions associated with respiratory syncytial virus and influenza virus infection in analyses applying broader inclusion criteria for prescription plan utilization.**

| | +/- 1-year Rx window* | +/- 2-year Rx window* |
|---|---|---|
| N antibiotic prescriptions | 18,466,530 | 18,466,530 |
| N person-weeks | 784,505,730 | 902,757,005 |
| Approximate antibiotic prescription incidence per 1000 person-weeks | 23.5 | 20.5 |
| Proportion of antibiotic use associated with RSV (95% confidence interval) | 6.3% (5.2-7.3%) | 6.0% (4.7-7.2%) |
| Proportion of antibiotic use associated with influenza (95% confidence interval) | 3.4% (3.1-3.8%) | 3.7% (3.3-4.1%) |

**Limited to study period of 2009-2017 to ensure capture of 2 years of prescription data for all cohort members.*

**Table S13. Incidence and proportion of outpatient antibiotic prescriptions associated with respiratory syncytial virus and influenza infection linked to diagnoses for which antibiotics are not indicated, without exclusion of prescriptions linked to other conditions, among children aged 0-17 years, 2008-2018.**

| **Diagnosis**[a] | **Antibiotic prescriptions among children 0-17 years** | | |
|---|---|---|---|
| | *Incidence rate, per 1000 person-years (95% CI)*[b] | *RSV-associated fraction, % (95% CI)* | *Influenza-associated fraction, % (95% CI)* |
| Bronchiolitis[c,d] | 9 (4, 16) | 24.2 (22.1, 26.1) | 4.0 (3.1, 4.9) |
| Bronchitis[d,e] | 61 (47, 77) | 10.8 (8.9, 12.6) | 6.0 (5.4, 6.7) |
| Nonsupp. OM[d] | 48 (35, 63) | 8.1 (6.8, 9.4) | 2.2 (1.7, 2.7) |
| Allergy[d] | 36 (25, 48) | 4.1 (3.0, 5.2) | 1.5 (1.1, 1.9) |
| Asthma[d] | 38 (26, 50) | 8.8 (7.4, 10.2) | 3.5 (3.0, 3.9) |
| Viral URI (excl. RSV and influenza)[d] | 199 (173, 228) | 10.1 (8.6, 11.5) | 6.7 (6.1, 7.2) |
| Any RSV diagnosis[d] | 3 (0, 7) | 49.8 (47.8, 51.7) | 7.9 (6.6, 9.1) |
| Any influenza diagnosis[d,f] | 9 (4, 15) | 15.1 (13.2, 16.9) | 92.0 (91.4, 92.6) |

Abbreviations: RSV – respiratory syncytial virus; CI – confidence interval; Non-supp. OM – nonsuppurative otitis media; URI – upper respiratory infection.

[a] Based on ICD-9-CM and ICD-10-CM codes (**Table S3**) assigned at the most recent outpatient visit within a 4-day window. More than one diagnosis can be associated with an antibiotic prescription.

[b] Obtained by fitting number of antibiotic prescriptions divided by person-years to a Poisson distribution, sampling 100,000 random draws, and obtaining the median, 2.5$^{th}$ and 97.5$^{th}$ percentile values.

[c] Excludes diagnosis codes for bronchiolitis due to RSV.

[d] Antibiotic prescriptions with concurrent diagnoses of urinary tract and other bacterial infections excluded.

[e] Excludes diagnosis codes for bronchitis due to RSV.

[f] Includes all diagnoses of influenza, including influenza and pneumonia and influenza and AOM.

**Figure S1. Predicted and observed antibiotic prescription counts**

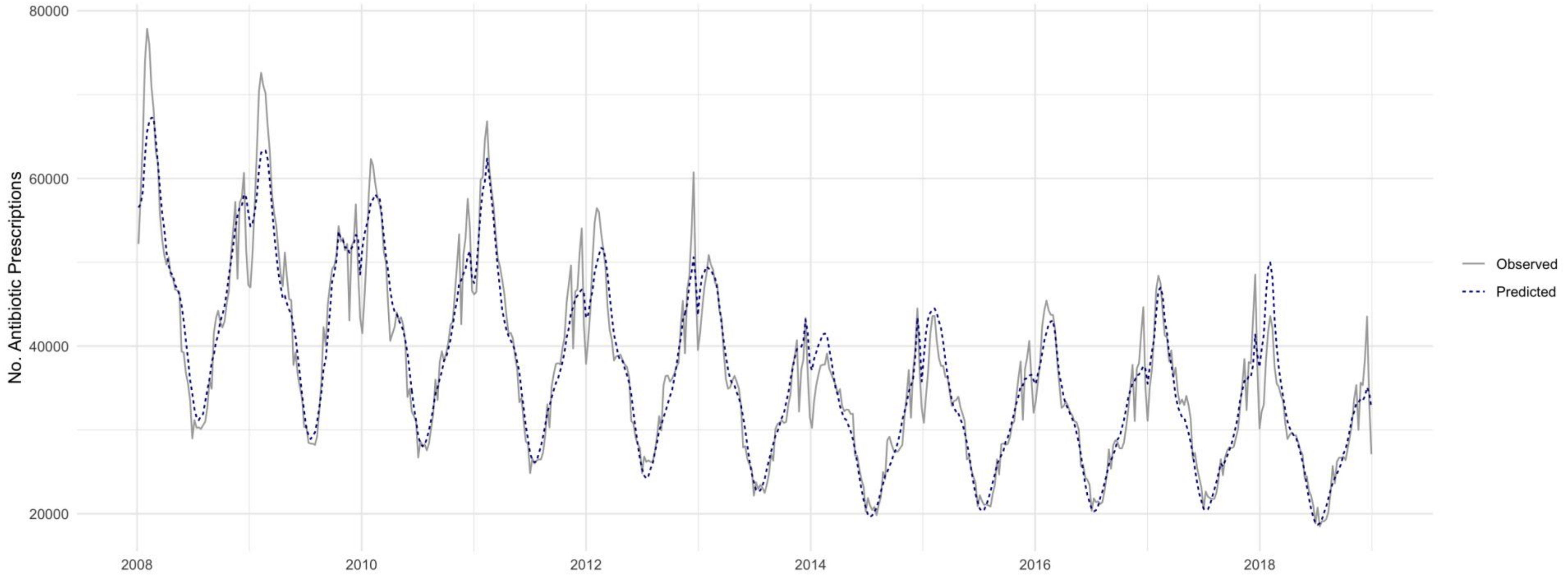
